# Genetics in Context: Social Factors Modify Polygenic Risk Across Multiple Cardiometabolic Diseases

**DOI:** 10.64898/2026.10.27.26366625

**Authors:** Micah R. Hysong, Sarah Hsu, Farida Akhtari, Miriam S. Udler, Josep M. Mercader, Alisa K. Manning, Michael D. Green, Iain R. Konigsberg, Luciana B. Vargas, Leslie Lange, Michael I. Love, Megan M. Shuey, LáShauntá M. Glover, Alison A. Motsinger-Reif, Genevieve L. Wojcik, Sara J. Cromer, Laura M. Raffield, Polygenic Risk Methods Development (PRIMED) Consortium

## Abstract

Polygenic risk scores (PRS) are increasingly used for disease prediction, but whether their associations vary across social contexts remains poorly understood. Here, we leveraged the large and diverse All of Us cohort to examine interactions between PRS and social determinants of health (SDoH) across eight prevalent chronic diseases and developed disease-specific polysocial risk scores (PsRS). We found consistent evidence of SDoH modification of PRS– disease associations for cardiometabolic diseases, including type 2 diabetes, coronary heart disease and chronic kidney disease, with PRS associations attenuated among more socially vulnerable populations. Joint modeling of PsRS and PRS significantly improved case status prediction across all eight diseases and PsRS were predictive for most diseases in external cohorts. These findings demonstrate that social context modifies associations between genetic risk and disease and suggest that integrating genetic and social information can refine disease prediction across diverse populations.

**Teaser:** Both genetics and the environment influence chronic disease, but whether and how they interact remains understudied. We find evidence of genetic effect modification of polygenic risk scores (PRS) by social factors for multiple cardiometabolic traits and that jointly modeling social risk improves disease prediction across a range of chronic diseases.

## Introduction

Chronic diseases, such as diabetes, heart disease, and chronic kidney disease, are influenced by myriad factors including genetics, lifestyle, and the environment; however, most research has assessed these risk factors independently.

Polygenic risk scores (PRS) estimate an individual’s genetic propensity for disease by aggregating the generally small effects of hundreds to thousands of genetic variants from population-based studies and are currently agnostic to lifestyle, environmental, or social context (1). PRS can capture a substantial proportion of population variance and therefore hold great promise for predictive and preventive medicine; however, their reduced accuracy in populations underrepresented in biomedical and genetic research stands as a major barrier to clinical implementation (2–5). While this poor transferability has been well documented for populations with increased genetic distance from the training sample, emerging evidence suggests that differences in the distribution of environmental and contextual factors such as smoking, income, and body mass index (BMI) may also contribute to disparities in cross-population PRS accuracy for certain traits (6–9). Moreover, environmental factors may modify associations between PRS and disease, complicating the interpretation and clinical application of PRS across the life course and diverse social and environmental contexts (5,10).

Early investigations into environmental modification of genetic effects have largely focused on behavioral and lifestyle factors which, while important, are at least partially modifiable at the individual level (11–17). In contrast, social determinants of health (SDoH) - including socioeconomic, sociocultural, healthcare access, and neighborhood or built environment features - significantly influence population-level health outcomes, are largely outside individual control, and are often upstream of behavioral and lifestyle factors (18) (19–22). Consequently, they are major systemic drivers of health disparities and inequities (22). Among these SDoH, socioeconomic status (SES) has been the most extensively studied in relation to genetic risk, largely due to the existence of commonly accepted, objective, quantifiable proxy measures and their relative availability in biomedical databases (23–25). Individual-level SES has been shown to impact PRS accuracy for quantitative traits at magnitudes comparable to genetic ancestry (6). Statistical interactions between PRS and individual- or area-level SDoH have been identified in studies of metabolic disease such as obesity and type 2 diabetes (T2D), whereas evidence for coronary heart disease (CHD) has been more limited, with weak interactions reported to date (26–29).

Despite these advances, existing studies are limited in three key ways: they have focused on a narrow set of traits (often studied independently in different contexts and with different analytical methods), considered a restricted range of SDoH (generally only including SES), and studied predominantly European-ancestry populations (26–28,30). Consequently, little is known about how genetic risk and social context jointly influence disease across diverse populations, social environments, and disease states. Failing to account for these interactions may lead to biased disease prediction, with disease being underestimated in socially disadvantaged populations and overestimated in socially advantaged populations, thereby exacerbating existing health disparities (6,31).

To address these gaps, we leveraged the *All of Us Research Program* (AoU), which is one of the largest and most diverse biomedical datasets with rich SDoH data. We systematically examined PRS effect modification by SDoH across eight chronic diseases prioritized by the electronic MEdical Records and GEnomics (eMERGE) Consortium for clinical implementation of PRS(32). We find consistent genetic effect modification for T2D, chronic kidney disease (CKD), and CHD by individual-level SES factors and area-level SDoH factors, as well as additional interactions with more extensive individual-level SDoH measures from SDoH surveys. Jointly modeling genetic and social risk improved case status prediction across all eight chronic diseases. Social risk scores, which aggregate multiple SDoH measures, also showed significant associations with four of six diseases evaluated in independent external healthcare cohorts, supporting the generalizability of social risk information across datasets. Together, these results advance equitable precision medicine by highlighting the importance and utility of integrating social context into genetic risk prediction models.

## Methods

### Cohorts and analytic variables

#### Main study and derivation cohort

The *All of Us Research Program (AoU)* is an NIH-funded initiative to increase the representation of groups historically and currently underrepresented in biomedical research within the United States (33). At the time of version release 8 (V8), AoU had 414,830 participants with whole genome sequencing (WGS) (34). Of those, 135,205 had sufficient electronic health records (EHRs) to classify disease status – defined as having at least three separate entries spanning at least three years. The final cohort of 104,946 participants also excluded those with incomplete SDoH measures described below (Supplementary Table 1; Supplementary Figure 1).

#### Validation cohorts

Mass General Brigham Biobank (MGBB) is a hospital-based biobank with more than 170,000 consented individuals linked with EHRs from the Mass General Brigham system based in Boston (35,36). 64,050 of these individuals also have genotyping data and 22,183 have complete survey information, zip code, and at least three years of records including at least three encounters (Supplementary Table 2). The MGBB cohort had a mean age of 63.51 (16.35) years, with 58.9% female, 89.8% White, 69.1% college-educated or with an advanced degree, and 61.2% privately insured participants.

The Personalized Environment and Genes Study (PEGS), an initiative of the National Institute of Environmental Health Sciences (NIEHS), includes nearly 20,000 North Carolina participants with multidimensional health and exposure data (37). WGS is available for 4,737 participants with the most complete survey data, of whom 4,265 had the required genotype, survey, and ZIP code data (Supplementary Table 3). The PEGS cohort had a mean age of 50.44 (± 14.84) years, with 68.4% female, 78.2% White, 15.4% Black or African American, and 67.3% college-educated or with an advanced degree.

#### Social Determinants of Health measures

Primary analyses used two individual-level (educational attainment and poverty) and seven area-level SDoH measures. Individual-level measures were derived from the AoU Basics Survey and selected based on prior evidence of predictive value and reduced selection bias(38). Income-to-poverty ratio (henceforth, “poverty”) was calculated as household income relative to the federal poverty threshold accounting for household size, location, and survey date and analyzed as a continuous variable; educational attainment was coded as a five-level ordinal measure. Area-level measures were derived from the American Community Survey 5-year estimates and aggregated at the 3-digit zip code level using population-weighted averages and temporally matched to AoU data, with values independently derived for consistency across cohorts. They included percent receiving government assistance, without a high school degree, living in poverty (<100% of the federal poverty limit), and without health insurance and community-level vacant housing, median household income, and a composite deprivation index(39).

A more comprehensive individual-level SDoH analysis used the AoU SDoH and Healthcare Access and Utilization surveys in a subset of 47,442 participants with more complete data; this sample has known response bias toward older, White, higher-income, and more highly educated individuals (Supplementary Tables 4-5)(38,40). Twenty-two SDoH variables, including perceived neighborhood crime, food insecurity, health coverage, and home ownership, were obtained from the AoU community workspace and derived as previously described(38,40). Missing data were multiply imputed using chained equations for participants with ≥60% survey completion. All SDoH measures were oriented such that higher values indicate greater social vulnerability.

### Genetic and clinical variables

#### Disease selection

Nine chronic diseases were initially selected based on prevalence, cost, and medical actionability in the eMERGE Network (32): asthma, atrial fibrillation (Afib), breast cancer, chronic kidney disease (CKD), coronary heart disease (CHD), hypercholesterolemia, prostate cancer, type 1 diabetes (T1D), and type 2 diabetes (T2D). Hypercholesterolemia was excluded because its PRS was inversely associated with case status before and after PC adjustment, leaving eight diseases for analysis.

#### PRS and PC adjustment

We used eMERGE-selected PRS, which were validated to be predictive in multiple ancestry groups(32). AoU-provided genetic PCs and genetic similarity groupings were used to PC-adjust PRS following Lennon et al. (2024): raw PRS were calibrated by modeling their mean and variance as functions of 16 genetic PCs and converting individual scores to PC-predicted z-scores (Supplementary Figures 2-3) (34); (32). The same procedure was applied to calibrate PRS in external validation cohorts. As PCs and SDoH are moderately correlated (average r ≈ 0.3 with PC1), this calibration substantially reduced the PRS-SDoH correlations (Supplementary Figures 4-5).

#### Disease ascertainment

Validated eMERGE algorithms were used to ascertain prevalent asthma, Afib, breast cancer, CKD, CHD, and prostate cancer, which are used in this study (32). AoU-specific algorithms were used for T1D and T2D (41). MGBB T1D and T2D algorithms were based off of the eMERGE algorithms, with modifications for diagnosis codes and diabetes medications to increase specificity. PEGS disease status was based on self-report because linked EHRs were unavailable. EHR algorithms used stringent control definitions to improve classification in the setting of incomplete records. Thus, case vs. control status reflects ascertainment of prevalent cases using these validated EHR-based phenotyping algorithms rather than the true population prevalence; case and control characteristics are provided in Supplementary Table 6.

#### Covariates

Primary models adjusted for age, age², sex/gender, record depth, and visit frequency. Sex/gender was categorized as cisgender female, cisgender male, or sexual and gender minority (SGM), with SGM excluded from CKD and T1D models because of insufficient case counts (<20); sex/gender was also omitted from breast and prostate cancer models. Age was defined at the most recent EHR encounter. Age^2^ was also included to control for non-linear associations between age and disease. Record depth was the number of unique encounters containing an observation or condition code, and visit frequency was record depth divided by EHR duration. Genetic PCs were included in sensitivity analyses but not primary models due to their correlation with SDoH; ancestry-related variation in PRS was instead addressed through PC-based mean–variance normalization to minimize potential over-adjustment (32).

### Statistical Analyses

#### SDoH-stratified analyses

For each of nine SDoH measures, participants were divided into quintiles, with Q5 indicating highest social vulnerability (Supplementary Figure 6; Supplementary Table 7). Tied values resulted in unequal quintile sizes (20,905–28,807; mean = 21,657.5, SD = 1,829.4) (Supplementary Table 8). For each disease–SDoH pair (8 diseases × 9 SDoH), logistic regression models were fit separately in Q1 and Q5 using the covariates described above and disease-specific PRS. Differences in PRS effects were assessed using two-sided z-tests. Sensitivity analyses additionally adjusted for genetic PCs, restricted analyses to African-like and European-like genetic similarity clusters, and downsampled the full cohort to match the sample size of the smaller African-like cluster.

The more comprehensive individual-level SDoH survey stratified analysis additionally controlled for individual-level poverty and education (Supplementary Figure 7; Supplementary Table 9).

Given the exploratory nature of these analyses, in the smaller and less diverse cohort, multiple testing was controlled using a more lenient Benjamini–Hochberg false discovery rate (FDR) correction.

#### Interaction analyses

For each disease–SDoH pair, logistic regression models included the covariates described above, PRS, SDoH, and a PRS × SDoH interaction; PRS and SDoH measures were z-score normalized. Conditional disease probabilities were estimated across the observed PRS range at SDoH levels corresponding to the 10th, 50th, and 90th percentiles, holding other covariates at typical values. Differences in PRS slopes between the low and high SDoH levels were tested using z-tests based on the interaction term. For both analyses, significance was Bonferroni-adjusted for 72 comparisons (6.94 × 10 ).

#### Net reclassification index

For each disease, we fit three nested logistic regression models: Base (age, age², sex/gender, record depth, and visit frequency), Genetic (Base + PRS), and Joint (Genetic + social risk score). NRI was calculated using the nri function in the mcca R package for Base-to-Genetic and Base-to-Joint comparisons, using both threshold-free predicted probabilities and 10% and 5% predicted-risk categories (42). Ninety-five percent confidence intervals were estimated from 100 nonparametric bootstrap samples.

### Construction and evaluation of disease-specific PsRS

#### Construction

For each disease, a disease-specific polysocial risk score (PsRS) was constructed using elastic net–regularized logistic regression (α = 0.5) incorporating individual poverty, individual education, and seven area-level SDoH measures. Five-fold stratified cross-validation was used, with λ selected by 10-fold cross-validation within each training fold; preprocessing was performed using training data only (Supplementary Figure 8) (43). Each PsRS was associated with increased probability of disease.

#### Evaluation

PsRS predictive value was evaluated for each of eight diseases using five-fold cross-validated logistic regression comparing five models: Base (demographic and EHR covariates), Social (Base + PsRS), Genetic (Base + PRS), Joint (Base + PRS + PsRS), and Interaction (Joint + PRS × PsRS). Performance was assessed using out-of-fold predictions, with bootstrap resampling (n=5000) used to assess statistical significance for paired AUC differences between the Genetic and Joint models. PsRS odds ratios were estimated from pooled out-of-fold predictions. Sensitivity analyses additionally adjusted Genetic, Joint, and Interaction models for 16 genetic PCs and replaced PsRS with individual SDoH measures.

#### Calibration

Calibration was evaluated for the Genetic and Joint models within each SDoH quintile. Observed-to-expected ratios were calculated from the sum of observed cases divided by the sum of the predicted probabilities; a ratio of 1 indicates agreement, values above 1 indicate underprediction, and values below 1 indicate overprediction. Expected Calibration Error (ECE) was calculated for each disease, model, and SDoH variable as the participant-count-weighted mean absolute difference between mean predicted risk and observed case proportion across quintiles.

#### Transportability

For transportability, disease-specific PsRS weights and scaling parameters were refit using the full AoU cohort and applied to MGBB and PEGS. Because MGBB lacked individual-level income/poverty data, an analogous PsRS excluding these measures was trained in AoU. The five models were evaluated in both external cohorts, excluding EHR-specific covariates from PEGS. PsRS associations were estimated from the Social models and combined across MGBB and PEGS using random-effects meta-analysis.

## Results

### Main study and derivation cohort description

Participants in the *All of Us Research Program* (AoU) with WGS, EHRs with at least three health care visits over three years, individual-level poverty and education data, and available three digit zip codes were included in this study (n=104,956; Supplementary Figure 1) (38). The resulting cohort had a mean age of 57.30 (± 16.49) years, with 60.3% cisgender female, 67.2% White, 11.4% Hispanic, 53.5% college-educated or with an advanced degree, 18.9% disabled, and 11.0% born outside of the United States (Supplementary Table 1). Based on genetic similarity to reference populations, 69.0% of participants aligned with European, 18.5% with African, 10.0% with Admixed American, and approximately 1% or fewer aligned with East Asian, Middle Eastern, or South Asian genetic ancestry (34).

### PRS and SDoH associate with disease

The number of prevalent cases ranged from 1,098 for T1D to 17,747 for T2D (Supplementary Table 6). In the overall AoU study cohort, each PRS, which was mean- and variance-normalized using genetic principal components (PCs), was significantly associated with AoU case status after adjustment for sex/gender, age, age², visit frequency, and record depth (Figure 1; top row). Effect sizes varied across diseases, ranging from 1.25 times the odds of asthma to 1.85 times the odds of T2D per one standard deviation (SD) increase in PRS (Supplementary Table 10).

**Figure 1:**
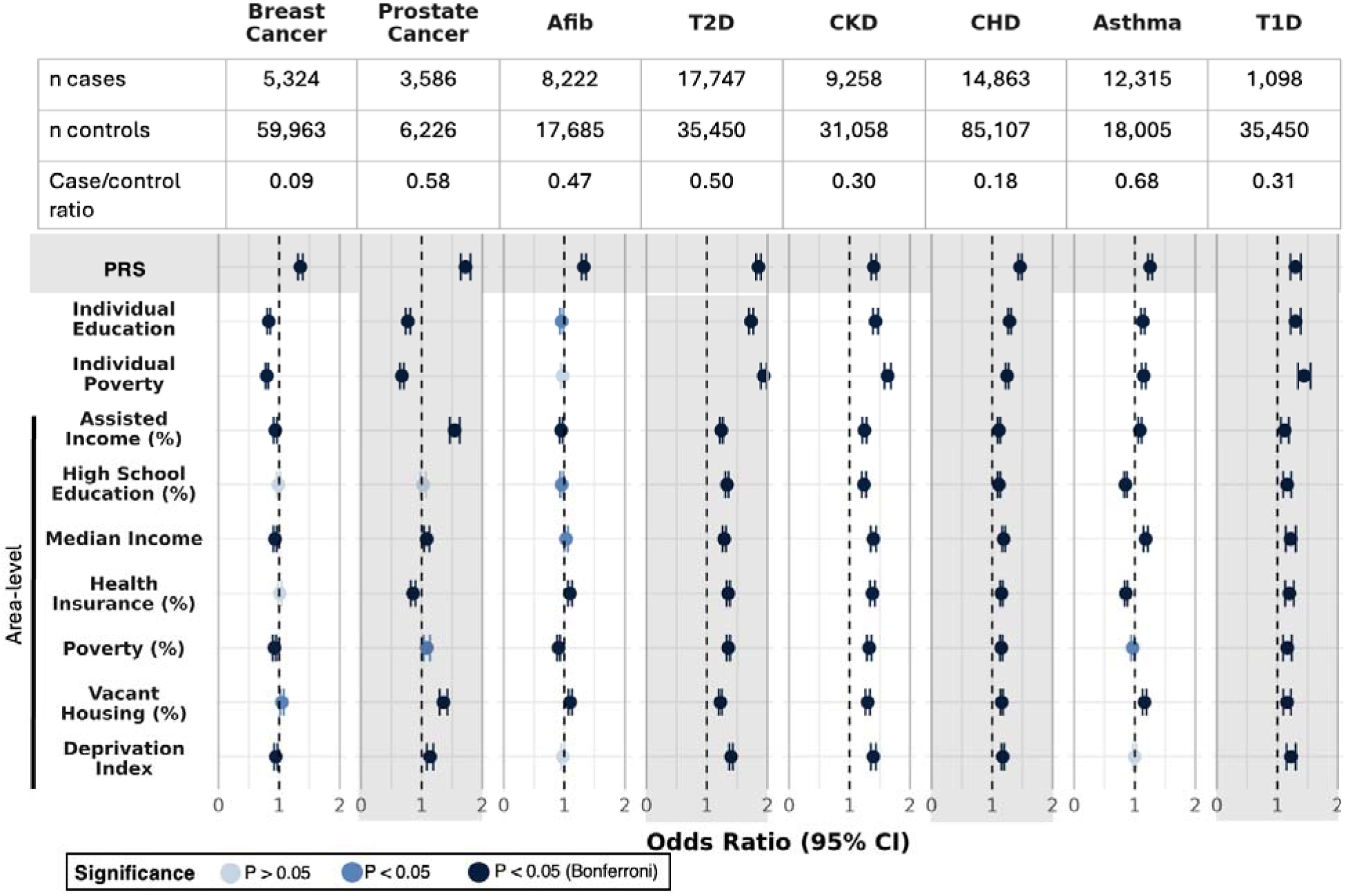
PRS and many SDoH are significantly associated with AoU case status. **Forest plots of odds ratios of PRS and SDoH for eight chronic diseases.** For each disease (columns) and SDoH variable (rows), the forest plots display the odds ratio (OR) and 95% confidence interval (CI) for PRS (top row; grey highlight) and SDoH variables. Dots are color coded by significance, where dark blue indicates statistical significance after Bonferroni correction (P < 6.25 × 10 ; 0.05/80 comparisons), blue indicates nominal significance p < 0.05, and light blue indicates p > 0.05. A null OR of 1 is indicated with a black vertical dashed line. Case sizes (in thousands) are displayed below each disease and the case/control ratio is displayed below that.

Of the 72 disease–SDoH pairs tested, 64 exhibited significant associations between SDoH and AoU case status after Bonferroni correction (P < 6.25 × 10 ). The direction of association was generally consistent across non-cancer diseases, with greater individual-level social vulnerability associated with higher odds of disease. In contrast, greater individual-level social vulnerability was associated with lower odds of breast and prostate cancer, potentially reflecting differences in disease ascertainment across social contexts (44). Notably, the association between individual-level education and disease was comparable in magnitude to that of PRS for T2D, CKD, and T1D, while associations with individual-level poverty were sometimes even larger. Adjustment for 16 genetic PCs as covariates attenuated some associations, which may reflect residual confounding by population structure (even with PRS whose means and variances were already adjusted for PCs) as well as shared variation between genetic ancestry and social context (Supplementary Figure 9).

### Genetic effect modification

To investigate whether the association between PRS and disease varies across levels of social vulnerability, we compared PRS–disease associations between individuals in the highest and lowest quintiles (Q1 vs Q5) of nine SDoH measures (Supplementary Table 8). We observed consistent evidence of effect modification for multiple cardiometabolic diseases, including T2D, CKD, and CHD, with PRS ORs differing significantly between SDoH extremes for most SDoH measures (Bonferroni-adjusted z test, P < 6.94 × 10 ) (Figure 2, Supplementary Table 11). For example, each one SD increase in PRS was associated with 2.15 [95% CI: 2.03, 2.27] times the odds of T2D in the lowest individual-level poverty quintile, compared with 1.58 [1.51, 1.65] times the odds in the highest poverty quintile (P=8.28 × 10 ¹ for the difference between quintiles). All non-cancer diseases exhibited evidence of effect modification by individual-level poverty. The association for Afib was only nominally significant, which may reflect the smaller number of cases and controls available for that analysis or the overall relatively weaker PRS association (OR 1.32) in this cohort. Across SDoH-trait pairs with significant effect modification, the relative effect of PRS on disease was always stronger in groups experiencing lower social vulnerability. Further reinforcing these findings, 18 of 19 SDoH–trait pairs (94.7%) that exhibited statistically significant differences in PRS ORs between SDoH extremes also had significant PRS x SDoH interaction terms in a whole population analysis not limited to SDoH extremes, with three additional pairs reaching significance (Supplementary Figure 10, Supplementary Table 12).

**Figure 2:**
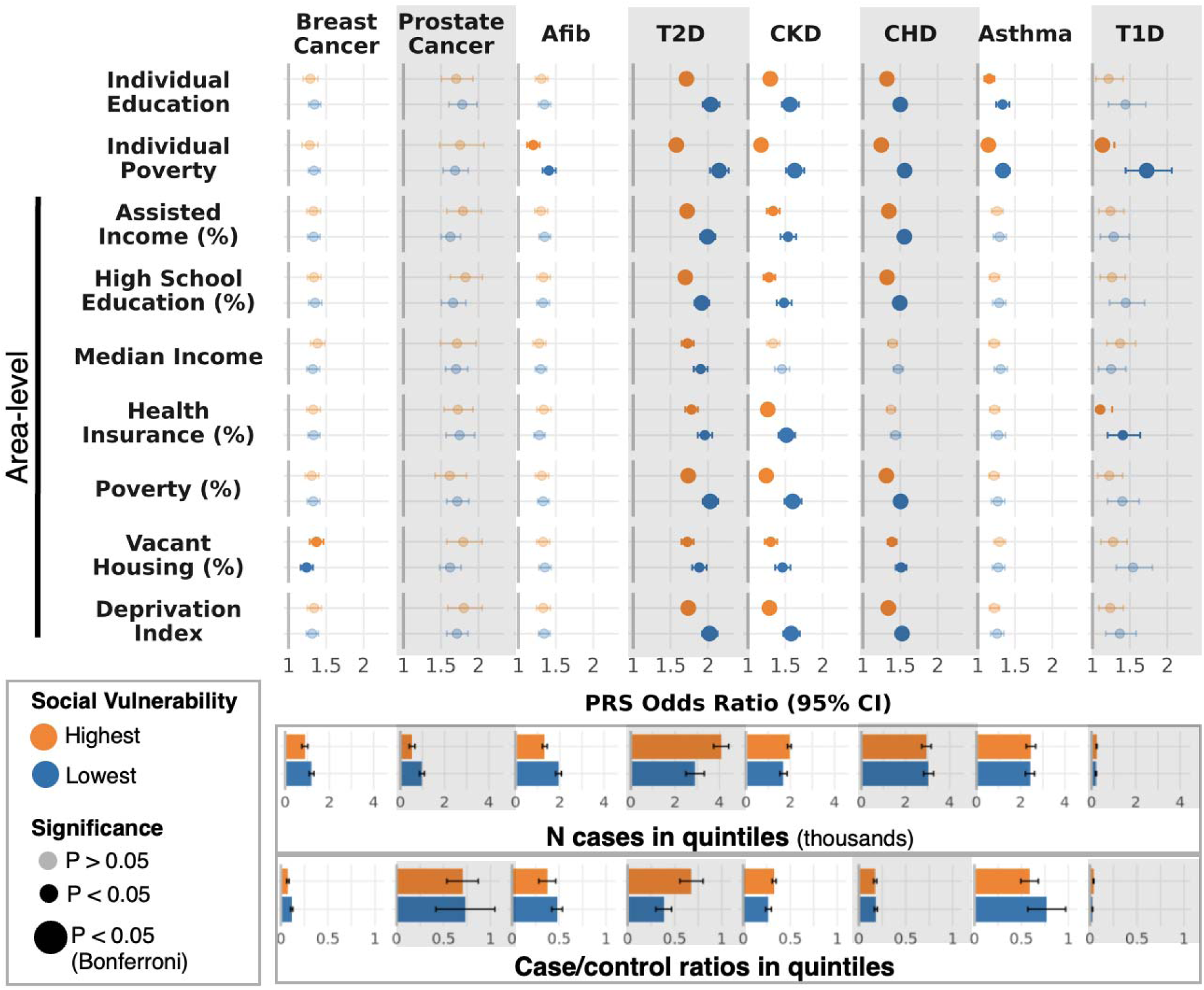
Larger effect of PRS in groups experiencing low social vulnerability for multiple cardiometabolic diseases. **Forest plots of odds ratios of PRS in SDoH stratified analyses.** For each disease (columns) and SDoH variable (rows), the forest plots display the odds ratio (OR) and 95% confidence interval (CI) for the PRS in the highest (Q5; orange) and lowest (Q1; blue) social adversity quintiles. Mean case sample sizes and case/control ratios for Q1 and Q5 across SDoH strata are displayed below each disease, with error bars representing ±1 standard deviation across SDoH strata. Case and control sample sizes for each individual disease–SDoH stratum are provided in Supplementary Table 6. Two-sided z-tests were used to assess differences between PRS ORs in the highest and lowest social adversity groups. ORs without significant differences between groups are displayed with increased transparency, whereas comparisons that remained statistically significant after Bonferroni correction (P < 6.94 × 10 ; 0.05/72 comparisons) are indicated by larger points.

Due to the complex population structure of the AoU cohort and reduced PRS accuracy in non-EUR genetic similarity groups (45), residual confounding may influence these findings if genetic ancestry proportions differ across SDoH strata, potentially contributing to differences in observed PRS–disease associations (Supplementary Figure 4). Therefore, to further assess potential confounding by population structure, we performed two sensitivity analyses. Additional inclusion of PCs as covariates led to largely consistent results (Supplementary Figure 11; Supplementary Table 13). We also repeated analyses within the two largest genetic similarity clusters (Supplementary Figure 12; Supplementary Table 14). Although this attenuated associations, a comparable loss of significance was observed in randomly downsampled whole-population analyses, suggesting that the reduction was largely attributable to decreased sample size rather than residual confounding by ancestry, though such tests should be repeated as sample sizes increase (Supplementary Figure 13; Supplementary Table 15). Therefore, we concluded that our results are likely not due to residual confounding by genetic substructure as captured by the PCs.

AoU also contains more comprehensive individual-level SDoH data derived from the SDoH and Healthcare Access and Utilization (HCAU) surveys. These measures were not included in the main analysis as requiring survey completion substantially reduced sample size and generalizability due to nonrandom response patterns that favor older, White participants with higher income and educational attainment, as described previously (Supplementary Table 5) (38,40). As such, these analyses are considered exploratory. To account for reduced sample size, this analysis was restricted to diseases with more than 3,000 cases among participants with available survey data (Afib, T2D, CKD, CHD, and asthma) (Supplementary Table 9). Even after controlling for individual-level poverty and education, we observed significant effect modification of the PRS by other SDoH factors following FDR correction, including food insecurity for Afib, CHD, and asthma; low social cohesion among neighbors for CHD; uninsured status for Afib and CKD; perceived neighborhood crime for CHD, and non-homeownership for Afib, T2D, CKD, and CHD (Figure 3; Supplementary Table 16). An additional fourteen pairs showed nominal significance (Supplementary Figure 14). Consistent with our primary analyses, PRS explained more variability in disease among socially advantaged groups across each SDoH domain. After additionally controlling for 16 genetic PCs as covariates, only food insecurity and perceived neighborhood crime remained associated with significant effect modification of the Afib and CHD PRSs (Supplementary Figure 15, Supplementary Table 17).

**Figure 3:**
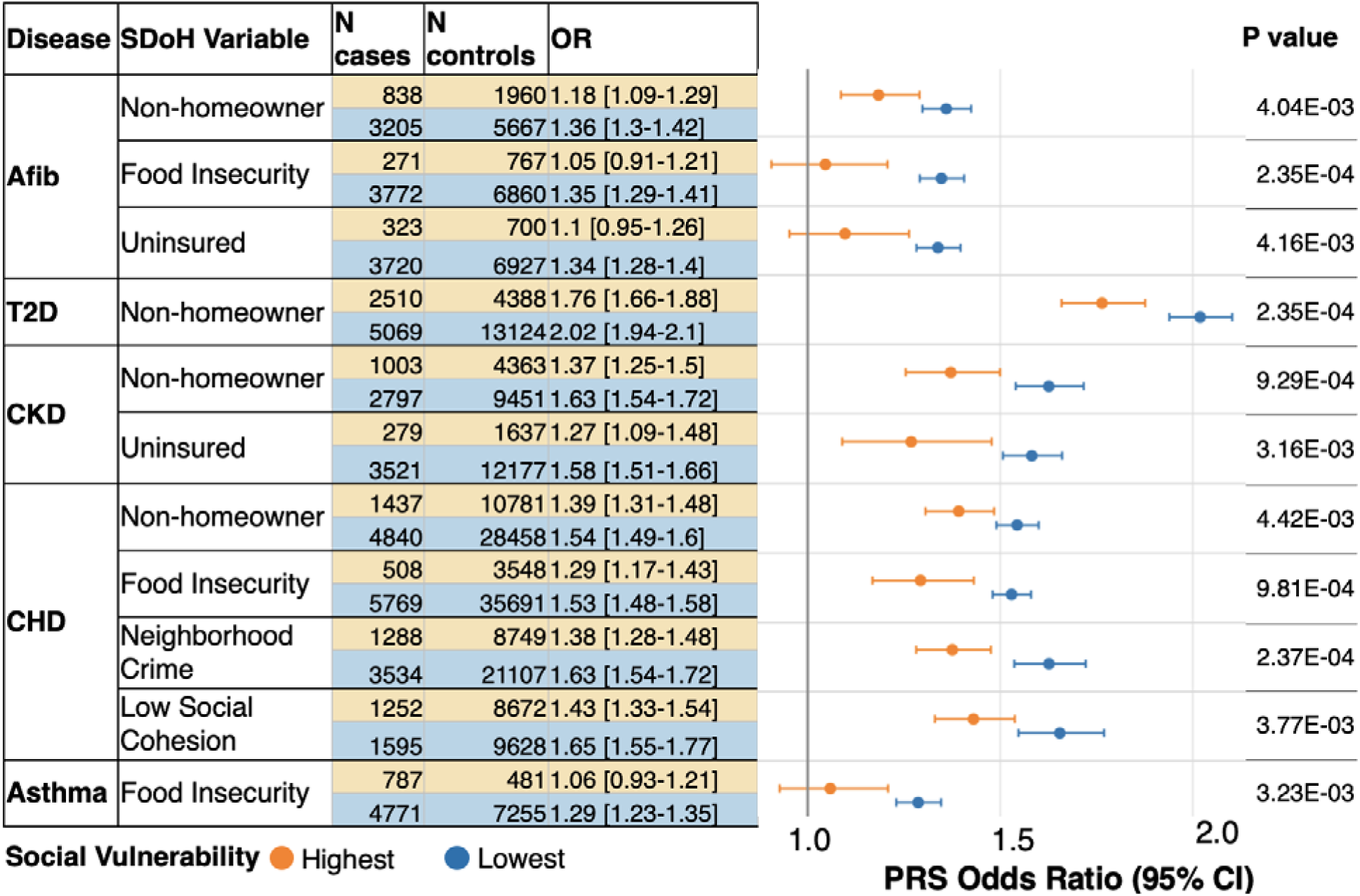
Larger PRS effects among groups with low social vulnerability for more comprehensive SDoH measures after adjustment for individual-level poverty and education. **PRS odds ratios in the lowest and highest individual-level SDoH quantiles for comparisons with significant differences after FDR correction.** Table and forest plot depicting PRS ORs for disease–SDoH pairs (from the more comprehensive individual-level SDoH survey) with significant differences between SDoH strata after FDR correction. Each disease–SDoH pair was evaluated independently in a separate model and statistical test. The table displays case and control sample sizes, ORs with 95% CIs, and P values from two-sided z-tests comparing PRS ORs between the highest (Q5; orange) and lowest (Q1; blue) social vulnerability groups. For binary SDoH variables, comparisons were made between lower and higher social vulnerability groups rather than quintiles. The forest plot displays the corresponding PRS ORs and 95% CIs, with point size proportional to the case sample size.

To better visualize the practical implications of these effect modifications, we plotted the conditional effects of PRS for an average individual at the 10th, 50th, and 90th percentiles of social vulnerability (Figure 4; Supplementary Figure 16). As expected, the predicted case probabilities increase with increasing PRS (Figure 4, x axis). Across diseases, predicted case probabilities are also higher among groups experiencing greater social vulnerability, particularly at lower levels of genetic risk (Figure 4, yellow > grey > blue lines). Across SDoH, we observe non-parallel/converging slopes, consistent with gene–environment interactions, wherein those with low genetic risk experience more variability in AoU case status by social environment.

**Figure 4:**
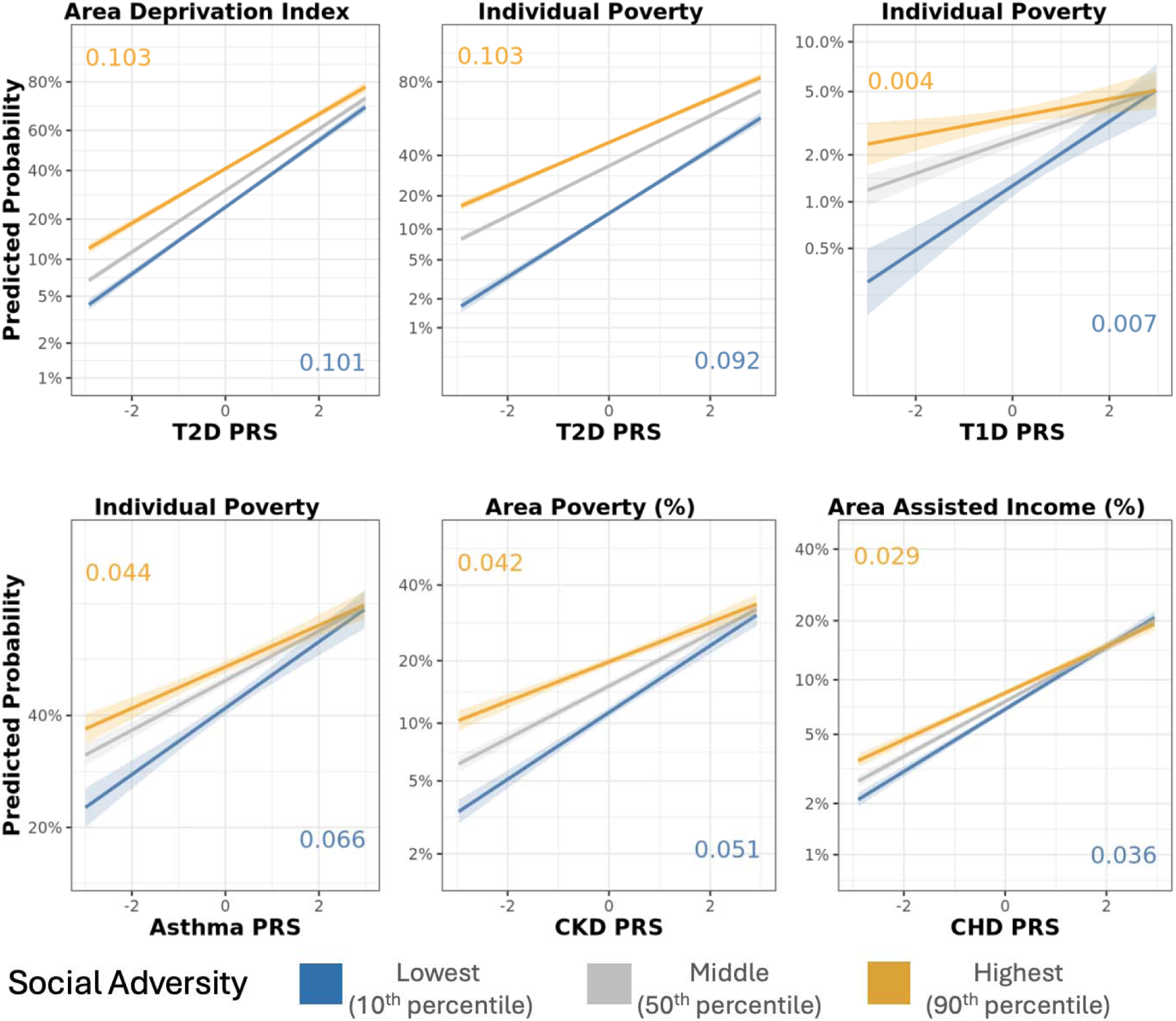
Social determinants modify polygenic risk across disease. **Effects plots for representative SDoH-disease pairs with significant interactions.** For each SDoH-disease pair, the PRS increases linearly across the x-axis, while the predicted probability of disease (based on case-control algorithms in AoU) is on a logit scale on the y-axis. Three effect curves represent low (10^th^ percentile; blue), intermediate (50^th^ percentile; grey), and high (90^th^ percentile; orange social vulnerability. Shaded areas denote 95% confidence intervals, with their slopes annotated in corresponding colors.

Alternatively, genetic risk has a greater effect on disease when social vulnerability is low (Supplementary Figure 17). For example, a one SD increase in PRS within participants at the 10th percentile of individual poverty increased the absolute probability of asthma case status by 7.2 percentage points, compared with 5.4 percentage points at the 90th percentile of individual poverty (Supplementary Tables 18-19; Supplementary Figures 17-18).

### SDoH explain heterogeneity within PRS-defined risk groups

Given that SDoH appeared to exert the greatest influence on AoU case status among individuals with low genetic risk, we next examined the extent to which SDoH explained AoU case status within the lowest genetic risk decile. To this end, and to optimally capture the contributions of both individual- and area-level SDoH, we constructed disease-specific polysocial risk scores (PsRS), in which elastic net models were trained independently on each disease to capture their unique social contributions from the nine different SDoH in the main, larger cohort (Supplementary Figure 8). As expected, we found that PsRS explained a large portion of case heterogeneity in the lowest PRS decile (Supplementary Figure 20; Supplementary Table 20). More relevant to clinical decision-making, PsRS also explained substantial heterogeneity in AoU case status within the highest PRS decile (Figure 5; Supplementary Tables 20-21). For example, among individuals in the top asthma PRS decile, case classification was approximately 30% lower in the lowest PsRS quintile and 50% higher in the highest PsRS quintile relative to the middle quintiles. Net improvement in reclassification showed a 7.4 percentage-point net improvement in continuous risk classification for prostate cancer, and more modest improvements for T2D and asthma (Supplementary Figures 21-22, Supplementary Table 22-24) (46).

**Figure 5:**
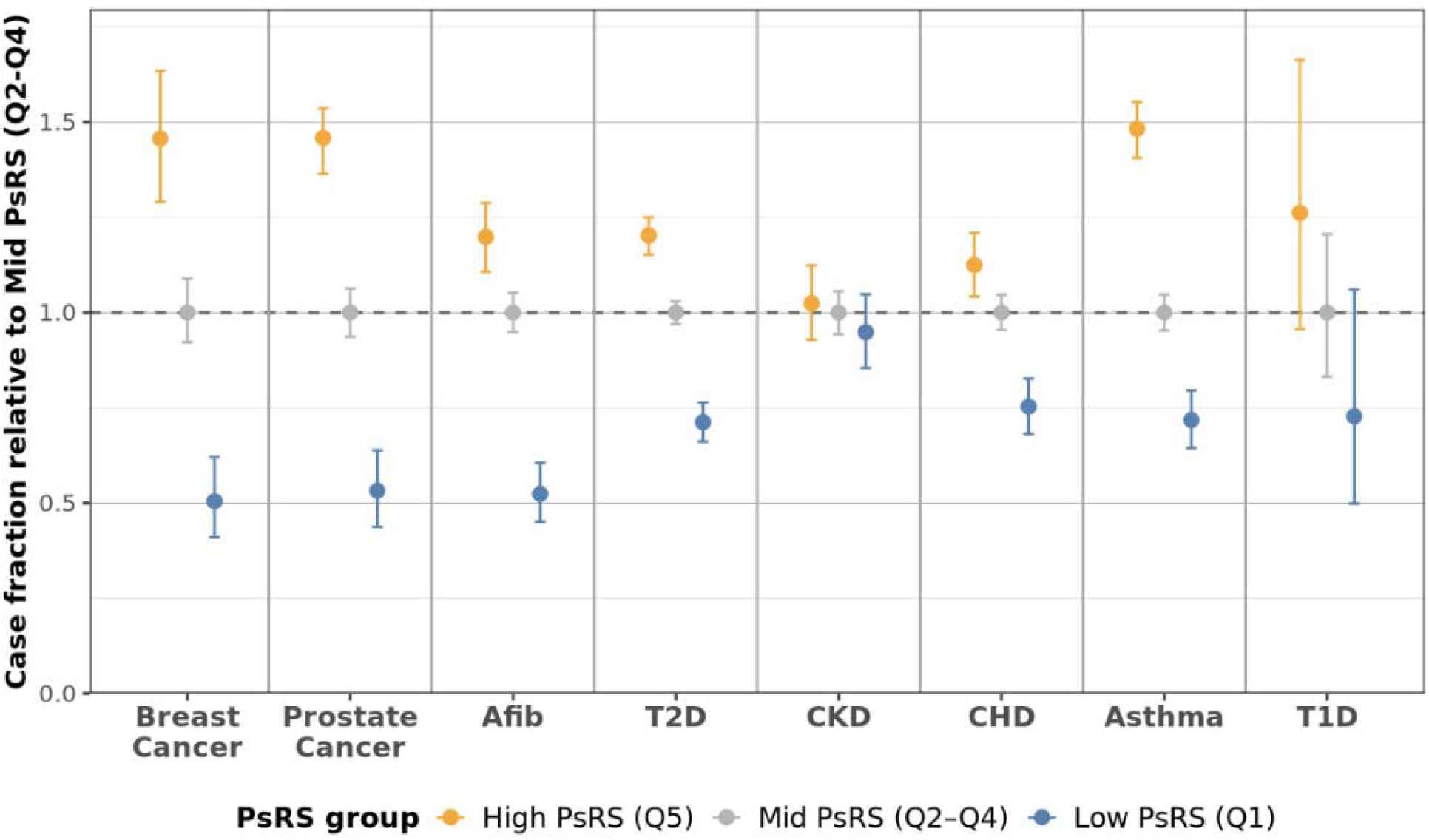
Substantial case status variation in top PRS decile by PsRS. **Dot plot of AoU case fraction among individuals in the highest genetic risk decile, stratified by PsRS quantile.** Case fractions are normalized within each disease to the middle PsRS group (Q2-Q4), such that values represent relative case fraction compared with the middle PsRS strata. These estimates reflect AoU case/control phenotype status, which uses stringent control criteria, and should not be interpreted as population-level disease prevalence. Error bars represent the 95% Wilson confidence interval for the observed case proportion, accounting for binomial sampling uncertainty. Blue dots represent low (Q1) PsRS, grey mid (Q2-4) PSRS, and orange represent high (Q5) social vulnerability.

### Combining genetic and social risk improves disease prediction modeling across cohorts

Next, we were interested in whether jointly modeling genetic and social risk factors could significantly improve disease prediction. While the magnitude of improvement varied, joint modeling PRS and PsRS significantly improved disease prediction for all eight diseases tested here (Figure 6; Supplementary Tables 25-26). For example, including PsRS in the joint model improved T2D AUC from 0.76 for the Genetic model to 0.80 (p<0.00020). Relative to the genetics model, the joint model also shrank the observed/expected ratio closer to 1 across SDoH quintiles for most SDoH-disease pairs and lowered the Expected Calibration Error (ECE) for all diseases except Afib and CHD (Supplementary Figures 23-24; Supplementary Tables 27-28). Despite significant interactions between PRS and SDoH with individual measures, including an interaction term between PRS and PsRS did not significantly improve AUC for any condition (Supplementary Figure 25; Supplementary Tables 29). Results remained significant in a sensitivity analysis adjusted for 16 genetic PCs (Supplementary Figure 26, Supplementary Tables 30-31).

**Figure 6:**
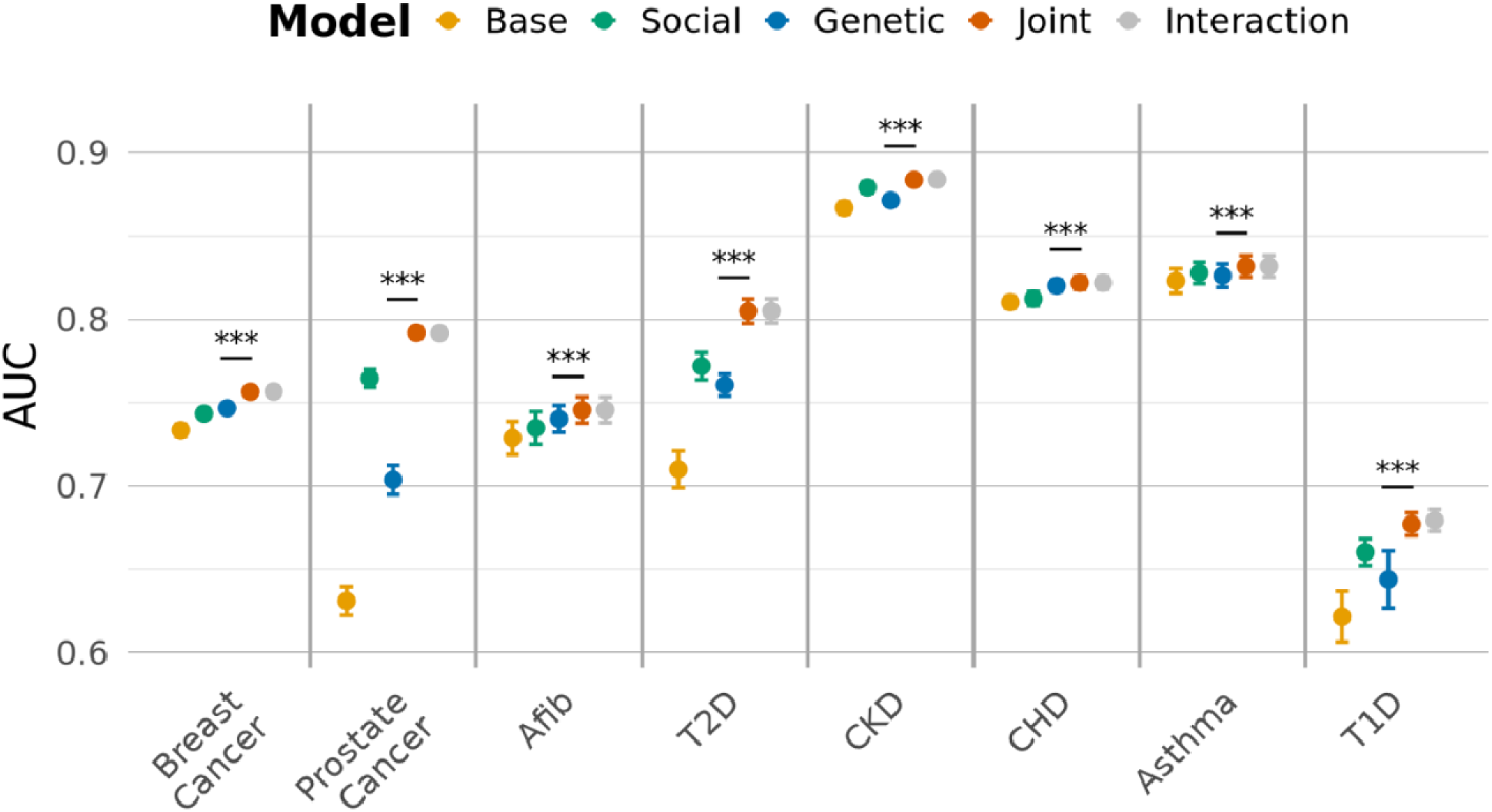
Joint modeling PRS and PsRS improves disease prediction. **Comparison of mean model discrimination (AUC) and SD across five-fold cross validation for eight diseases.** The **Base** model (orange) includes demographic and data availability covariates (age, age², sex/gender [except for breast and prostate cancer], record depth, and visit frequency). The **Social** model (green) extends the Base model by adding PsRS, while the **Genetic** mode (blue) adds PRS. The **Joint** model (vermillion) includes both PRS and PsRS, and the **Interaction** model (grey) additionally includes a PRS × PsRS interaction term. Significance comparisons are made between the Genetic and Joint model with 5000 bootstrap resamples. Three stars indicate p < 0.00020 (5000 bootstrap resolution).

To evaluate the generalizability of PsRS across healthcare systems and biobanks, we applied PsRS derived from a full-data refit in AoU to the Mass General Brigham Biobank (MGBB) and the Personalized Environment and Gene Study (PEGS). Afib and T1D did not have enough cases to be evaluated in these external cohorts. PsRS trained in AoU were significantly associated with disease odds for four out of six of the remaining diseases under the fixed-effects meta-analysis framework, indicating significant transferability (Supplementary Figure 27; Supplementary Table 32). Additionally, CHD and T2D PsRS were significant under the random-effects framework and showed the strongest transferability. For T2D, the random-effects meta-analysis estimated an OR of 1.50 [95% CI: 1.21, 1.85] per one SD increase in PsRS, compared with an OR of 2.01 [95% CI: 1.96, 2.05] in the AoU training cohort under the five-fold cross-validation framework, while the CHD PsRS demonstrated a similar association in the external and AoU cohorts.

PRS were more consistently and more strongly associated with disease in external testing cohorts (Figure 7). This was not always the case in AoU, indicating some overfitting of the PsRS AoU model. In MGBB, joint modeling PsRS alongside PRS and base covariates resulted in a statistically significant increase in AUC compared to the genetic model for every disease except asthma (Supplementary Figure 28; Supplementary Tables 33-34). Further demonstrating generalizability, PsRS trained in MGBB or PEGS performed similarly to those trained in AoU within their respective cohorts, although breast and prostate cancer sample sizes in PEGS were too small to train PsRS (Supplementary Figure 29).

**Figure 7:**
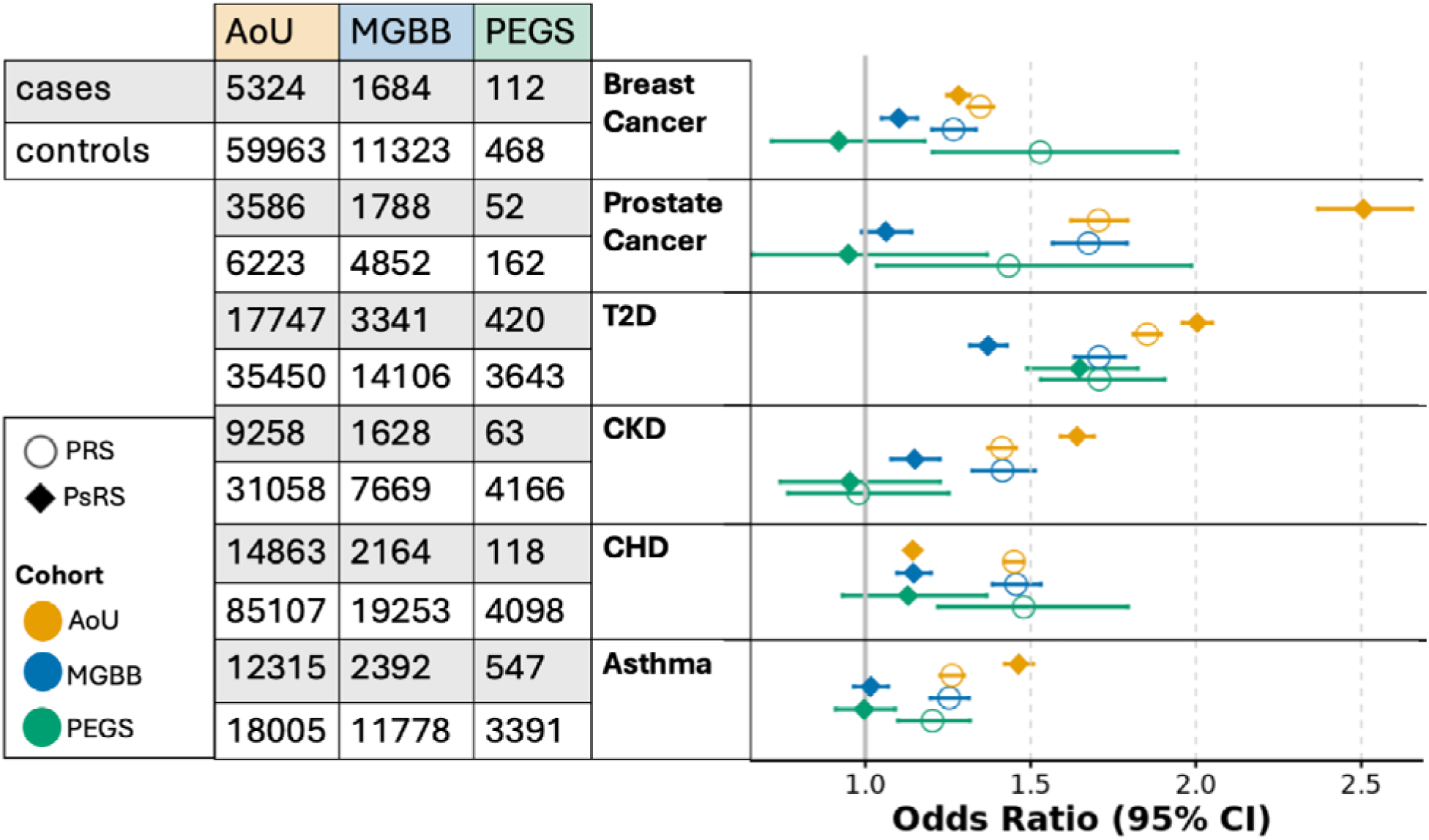
Effect estimates of PRS and AoU-trained PsRS across cohorts. **Forest plot of PRS and PsRS OR in AoU and external testing cohorts.** Odds ratios represent the change in disease odds per 1-SD increase in PRS or PsRS. Numbers of cases and controls are provided for each cohort. Point color denotes cohort, and point shape distinguishes PRS from PsRS.

## Discussion

We leveraged SDoH, genetic, and EHR data from the large, diverse AoU dataset to identify genetic effect modification by SDoH across multiple cardiometabolic diseases and develop transferable PsRS. When evaluating PRS associations for eight chronic diseases prioritized for clinical implementation, we found evidence of significant genetic effect modification by SDoH. PRS associations with cardiometabolic diseases were attenuated in socially vulnerable populations; social factors had a larger effect on AoU case status among individuals with lower genetic risk and a lesser effect among high genetic risk individuals. Nevertheless, social risk explained substantial case variation among individuals even in the highest decile of predicted genetic risk. Encouragingly, joint modeling of social risk, in the form of PsRS, alongside PRS significantly improved disease prediction for all eight diseases. Moreover, four out of six of the PsRS trained in AoU showed transferability in external datasets. Together these results highlight the power and utility of incorporating SDoH into disease prediction models alongside PRS.

In models stratified into top and bottom SDoH quintiles, we saw evidence of genetic effect modification by both individual and area-level SDoH for T2D, CKD, and CHD PRS, and by individual poverty for asthma and T1D PRS. Notably, PRSs with significant effect modification were those which had the strongest residual correlation with SDoH after PC mean and variance adjustment; however, the persistence of interaction effects after more extensive PC adjustment (additionally including PCs as covariates) indicates that these findings likely reflect, at least in part, genuine heterogeneity in PRS associations across social environments. This is further supported by PRS effect modification by SES in European-ancestry-only cohorts (47).

Moreover, all diseases with sufficient sample sizes to evaluate the more comprehensive individual-level survey measures (asthma, Afib, T2D CKD, and CHD), displayed effect modification by at least one other SDoH variable following FDR correction, even after controlling for individual poverty and education. While the SDoH survey–based analyses should be considered exploratory until these measures are more broadly collected in diverse populations (38), the findings suggest that individual-level SDoH beyond more commonly reported income and education SES metrics, for example healthcare coverage and home ownership, may be important to consider when evaluating PRS transferability and equitable clinical implementation.

For SDoH–trait pairs with significant differences in PRS odds ratios between the most and least socially advantaged groups, PRS consistently had a stronger association in the socially advantaged population, suggesting that environmental factors may contribute to lack of PRS transferability across populations with different social exposures (8,9,48). Negative interaction terms and conditional effect analyses indicated that SDoH had a stronger association with AoU case status among individuals with low genetic risk. In other words, as genetic risk increased, the association between social vulnerability and disease was attenuated; likewise, as social vulnerability increased, the predictive contribution of genetics was attenuated. One possible explanation is that when social factors account for a greater proportion of variation in disease risk, PRS may explain a smaller proportion of the remaining variation in disease risk (8,9,48).

This attenuation may also partly reflect differences in disease prevalence across social strata: when socially vulnerable groups have a higher underlying disease prevalence, a similar increment in genetic liability may correspond to a smaller relative increase in prevalence, resulting in an apparently weaker PRS effect on the multiplicative scale (47). An alternative explanation is that PRS are less predictive in socially disadvantaged populations due to their underrepresentation in Genome-Wide Association Study (GWAS) training data, although SNP effect sizes have been found to be relatively stable across environmental strata and generally each SNP explains a small portion of trait variance (30,49).

Considering SDoH, in the form of PsRS, alongside genetic risk improved disease prediction for all eight chronic diseases tested here. To evaluate their generalizability, we validated the PsRS in two independent cohorts, MGBB and PEGS, where they demonstrated moderate transferability. While PRS had stronger effects than PsRS in these cohorts, the PRS had more predictors and were trained on larger sample sizes than AoU alone. Moreover, each external cohort presented distinct limitations for PsRS construction (absence of income data in MGBB, small sample size in PEGS). Despite these limitations, fixed-effects meta-analyses of AoU-trained PsRS were significantly associated with disease odds for four of the six diseases, and T2D and CHD PsRS demonstrated the greatest transferability.

This study has several strengths. First, AoU is one of the largest and most diverse biomedical datasets and is among the first to collect genomic data, EHRs, and rich individual-level and area-level SDoH data. Second, we focused on clinically relevant chronic diseases and corresponding PRS already developed and validated by eMERGE, increasing translational relevance. Third, to our knowledge, this is one of the first studies to systematically evaluate interactions between PRS and SDoH across multiple chronic diseases within a single cohort, improving comparability across diseases and demonstrating the potential generality of these effects. Methodologically, we supported our findings using multiple complementary approaches—combining stratified analyses with formal interaction models—and ran sensitivity analyses to help address potential confounding from population stratification. We further extended prior work by incorporating a broader and more granular set of SDoH measures than is typical in PRS studies, enabling a more comprehensive assessment of social context. Finally, we developed PsRS that leverage the scale and diversity of AoU, and demonstrated their ability to improve disease prediction across a range of diseases and their transferability in external cohorts, supporting their potential real-world applicability in diverse healthcare settings.

This study also has some limitations. First, EHR completeness and depth, and SDoH survey data are subject to selection and data availability bias influenced by demographic, socioeconomic, and health care access factors ((38,40)). EHR-derived phenotypes are affected by clinical ascertainment, including screening intensity (e.g., for breast and prostate cancer), which may introduce residual confounding when interpreting gene–environment interaction effects. Second, detecting interaction effects typically requires substantially larger sample sizes than main effects. Accordingly, the presence of effect modification in higher-powered diseases and the absence of detectable interactions in others may partly reflect limited statistical power rather than true absence of effect modification. Third, PRS may capture population-stratification-related signal, particularly when genetic ancestry is correlated with social and environmental variables and when genome-wide construction methods are used (50). The eMERGE-selected PRS used in this study included both genome-wide (asthma, CKD, CHD, T2D) and sparse scores (breast cancer, prostate cancer, afib, T1D), potentially contributing to heterogeneity in the extent of population-stratification-related confounding across diseases. Due to the correlation between genetic PCs and social and environmental variables, adjustment for genetic PCs to reduce confounding from population stratification may also attenuate some true SDoH-related signal, thus providing a conservative estimate of interaction effects. Finally, the external validation results likely represent conservative estimates of PsRS performance due to the lack of individual-level poverty data in MGBB and the small sample size of the PEGS cohort.

As AoU continues to expand and other large-scale datasets increasingly incorporate SDoH measures, these findings should be replicated in independent and more diverse populations, as well as in traditional cohorts or settings with universal healthcare that will be less impacted by and biased according to healthcare access. In addition, as longer follow-up enables greater accrual of incident cases and mortality outcomes, future work should assess effect modification in incident disease contexts and for disease complications. Given known real-world biases in AoU, further studies should also evaluate the extent to which these factors influence the transferability of PsRS, particularly to ensure that they do not reinforce or amplify existing disparities. Moreover, future research should aim to disentangle genetic effect modification due to bias in the training data (for example higher rates of case control misspecification or underrepresentation of socially disadvantaged populations) from true gene-by-environment interactions. Finally, robust incorporation of SDoH into genetic and clinical risk models will require coordinated efforts across biobanks and healthcare systems to improve the collection, harmonization, and analytic integration of social determinants within disease prediction frameworks.

This work demonstrates the importance, power, and utility of integrating SDoH into genetic risk prediction models. Failing to account for social context may result in biased disease prediction where disease is underestimated in socially disadvantaged groups due to attenuated genetic effects and omission of key structural determinants of health. Although we focused on eight chronic diseases, complementary work suggests these patterns may generalize across a broader range of PRSs (51). As biomedical research increasingly incorporates social determinants at scale, future research should more systematically investigate these effects and develop robust, transferable social risk models. More broadly, these findings reinforce that personalized medicine requires moving beyond genetic risk alone. Integrating SDoH alongside PRS could offer a more complete and equitable framework for disease risk prediction and clinical implementation.

## Data Availability

Individual-level data for All of Us is available through the Curated Data Repository in the All of Us Researcher Workbench. Individual-level data for MGBB and PEGS cannot be shared because patient participants have not been broadly consented for dissemination of individual-level data.

https://workbench.researchallofus.org/login

## Code Availability

All modeling was performed in R (version 4.5.0) and analysis scripts are publicly available at https://github.com/micah-r-hysong/PRS_SDoH_AoU/. PsRS weights and scaling files and scripts used for external data validation, including disease algorithms, are available at https://github.com/micah-r-hysong/PRS_SDoH_analysis.

## Acknowledgments

The authors thank the participants and research teams from the All of Us research program, without whom this research would not have been possible. We would additionally like to thank the participants and research teams from PEGS and MGBB. We would also like to thank the Polygenic Risk Methods Development (PRIMED) Consortium Social Determinants of Health Working Group. A list of individuals included in PRIMED consortium banner are at https://primedconsortium.org/publications/banner.

