## Supplemental Figures for "Genetics in Context: Social Factors Modify Polygenic Risk Across Multiple Cardiometabolic Diseases"

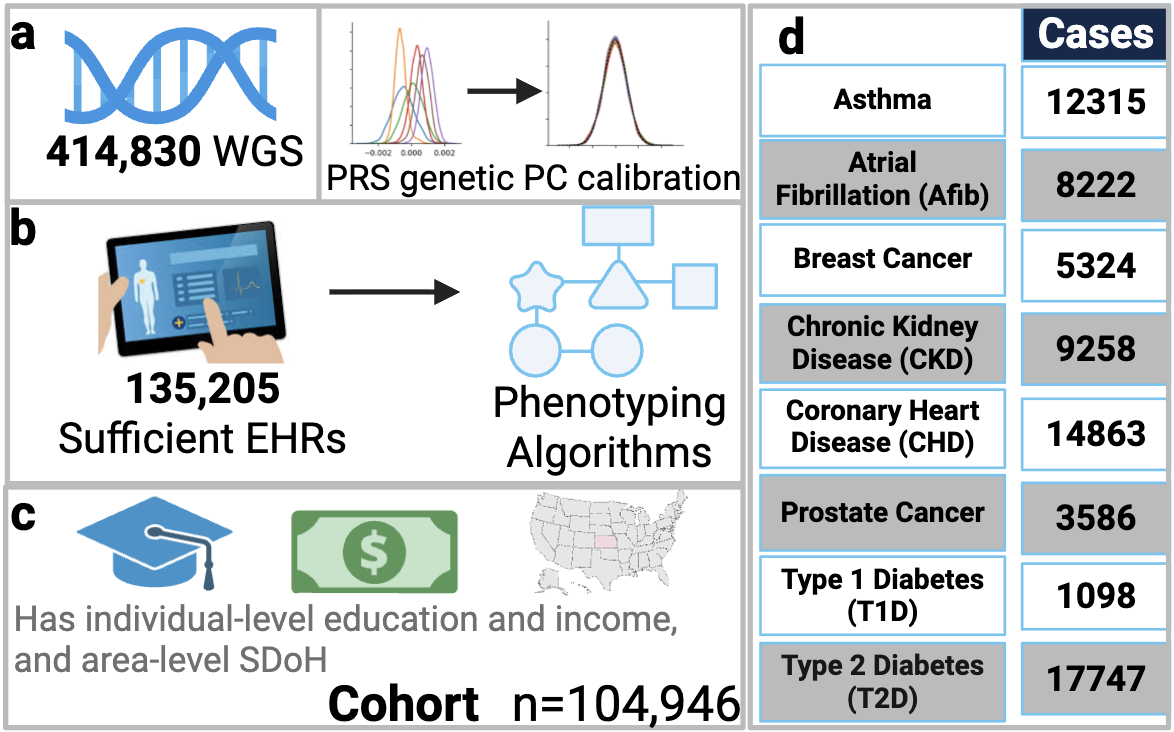

**Supplementary Figure 1**: Participant inclusion flow chart for All of Us. **A)** At the time of version release 8 (V8), 414,830 participants had Whole Genome Sequencing (WGS). Mean and variance Z-score normalization was performed on PRS generated from WGS. **B)** Of those with WGS, 135,205 had sufficient electronic health records (EHRs). Phenotyping algorithms were performed on these EHRs. **C)** Finally, 104,946 participants also had income, education, and area-level SDoH. **D)** Total number of cases for each disease.

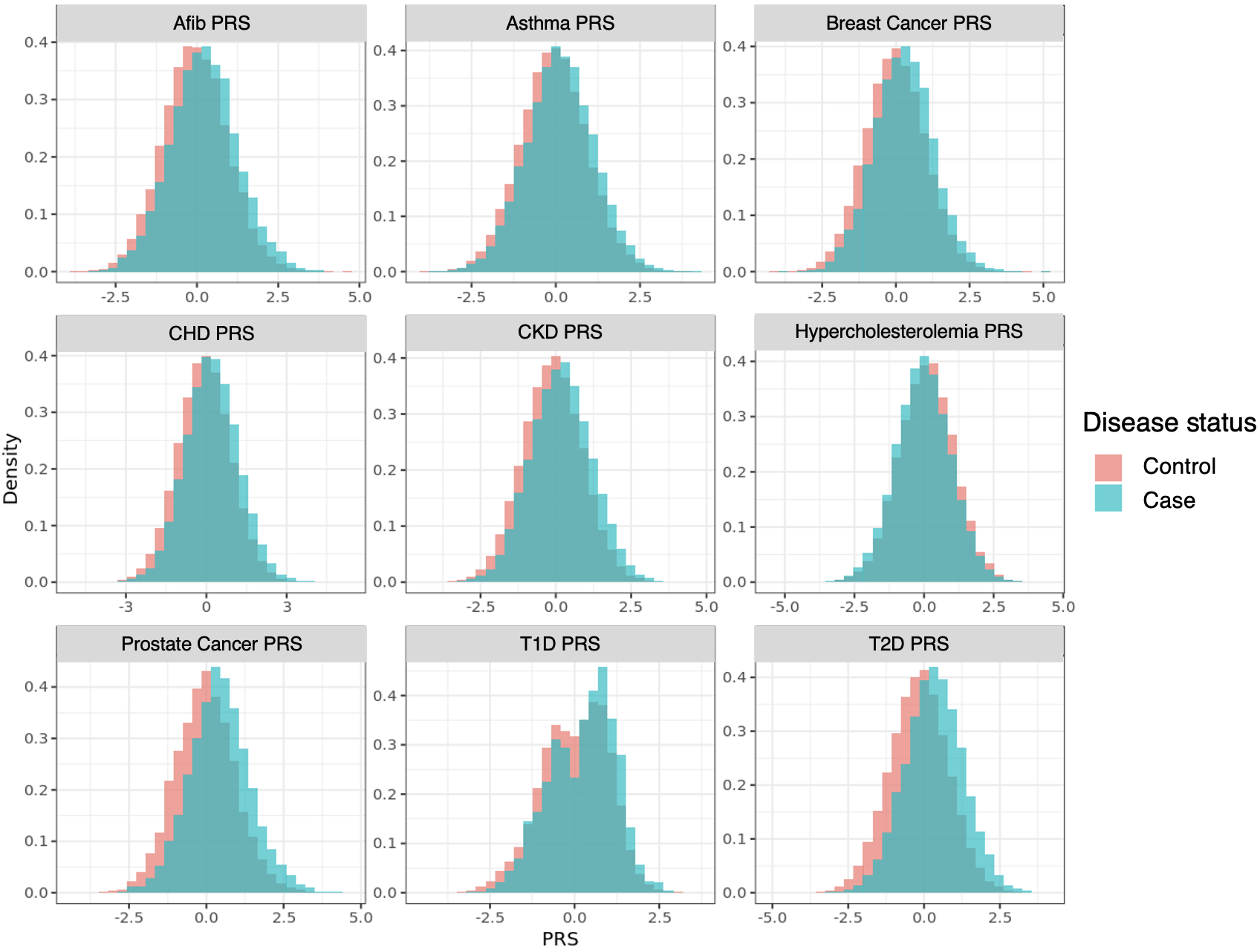

**Supplementary Figure 2:** Distribution of genetic PC normalized PRS by Case/Control status

Density plots of PRS after PC adjustment stratified by case (blue) vs. control (orange) status for nine chronic diseases.

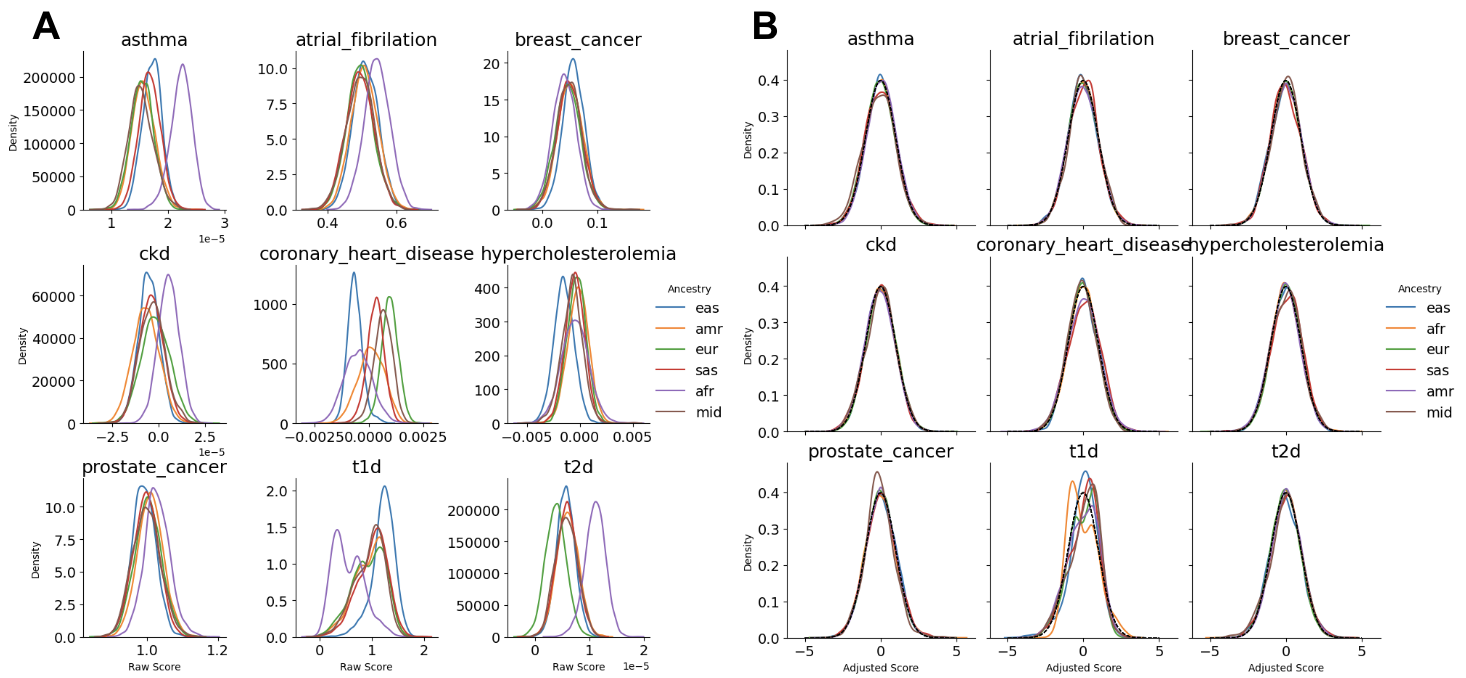

**Supplementary Figure 3:** PRS distributions **(A)** pre- and **(B)** post-adjustment

Density plots of PRS **A)** before and **B)** after mean and variance normalization using 16 genetic PCs. Curves are color coded by genetically inferred continental ancestry groups to demonstrate mean and variance shifts; however, groupings were not used in the adjustment.

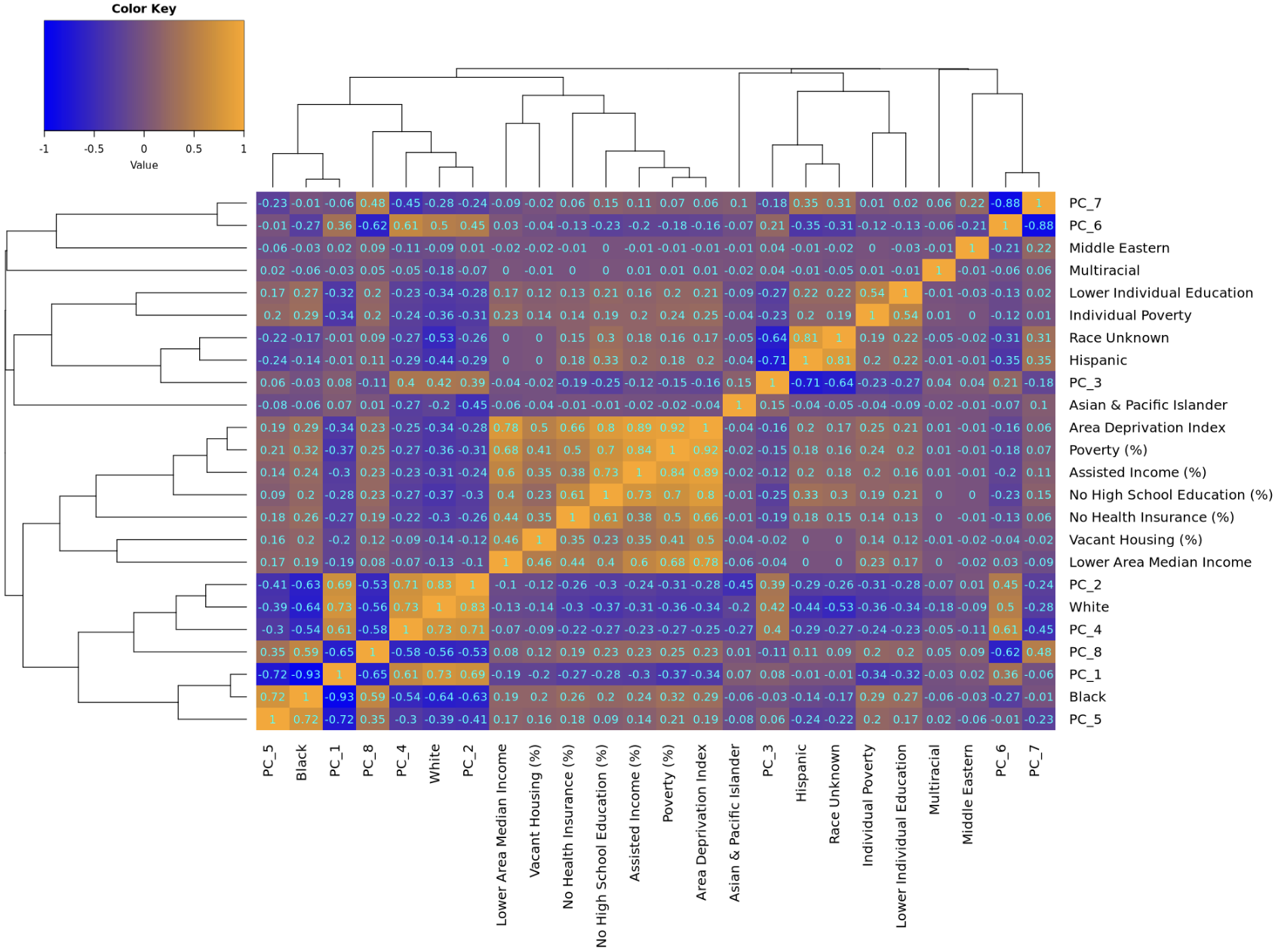

**Supplementary Figure 4:** Self-identified race and ethnicity (SIRE), Genetic Similarity PCs, & SDoH Correlation Heatmap. Correlation Heatmap between self-identified race and ethnicity (SIRE), genetic similarity to 1000G/HGDP reference populations principle components, and SDoH in the main cohort. Variables are arranged according to hierarchical clustering.

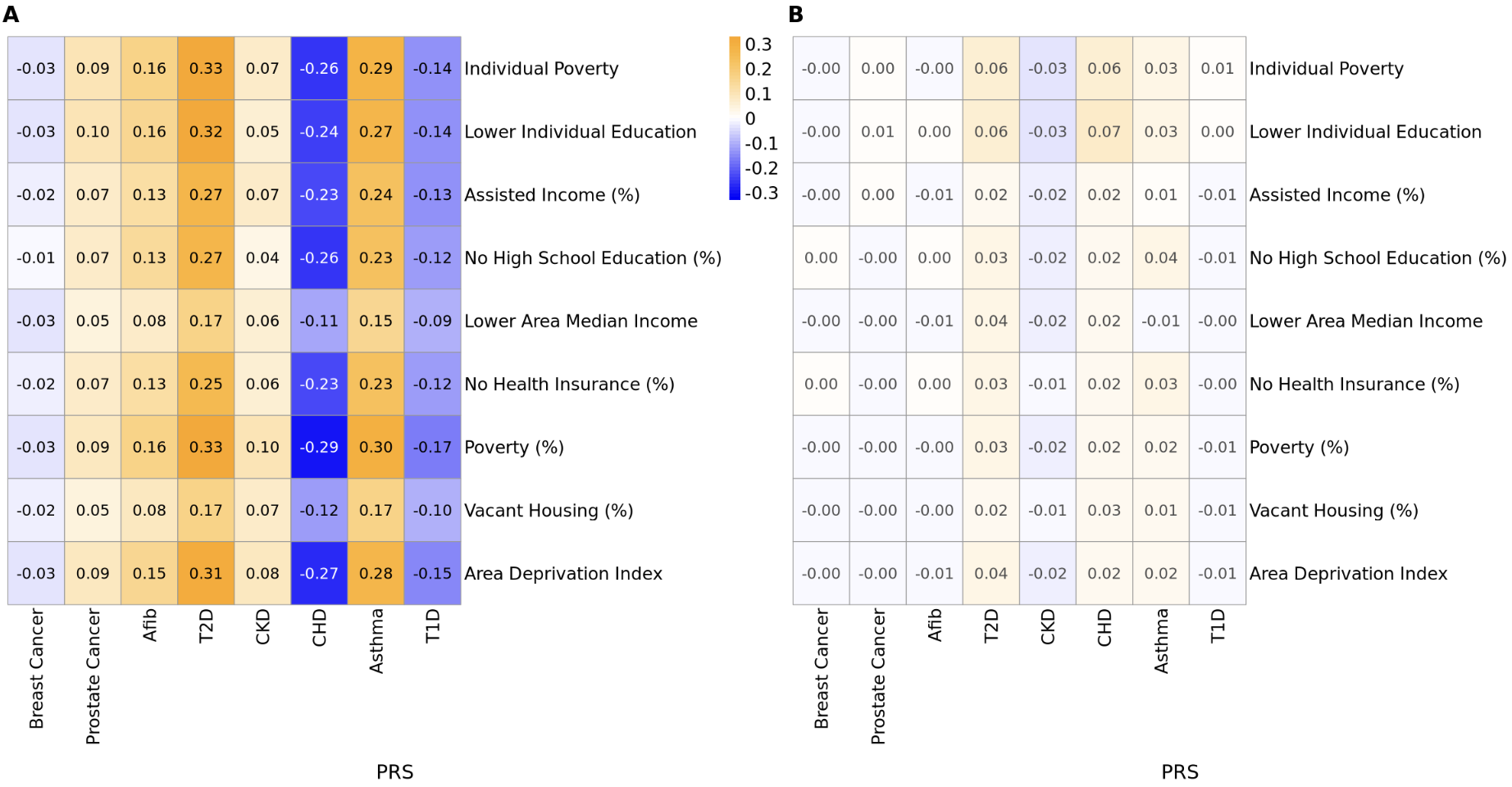

**Supplementary Figure 5:** Correlation Between SDoH and PRS **A)** before and **B)** after PC adjustment. Correlation heatmaps between SDoH and PRS **A)** before and **B)** after the PRS were adjusted for 16 genetic PCs. Heatmaps in A and B are on the same scale to facilitate comparison.

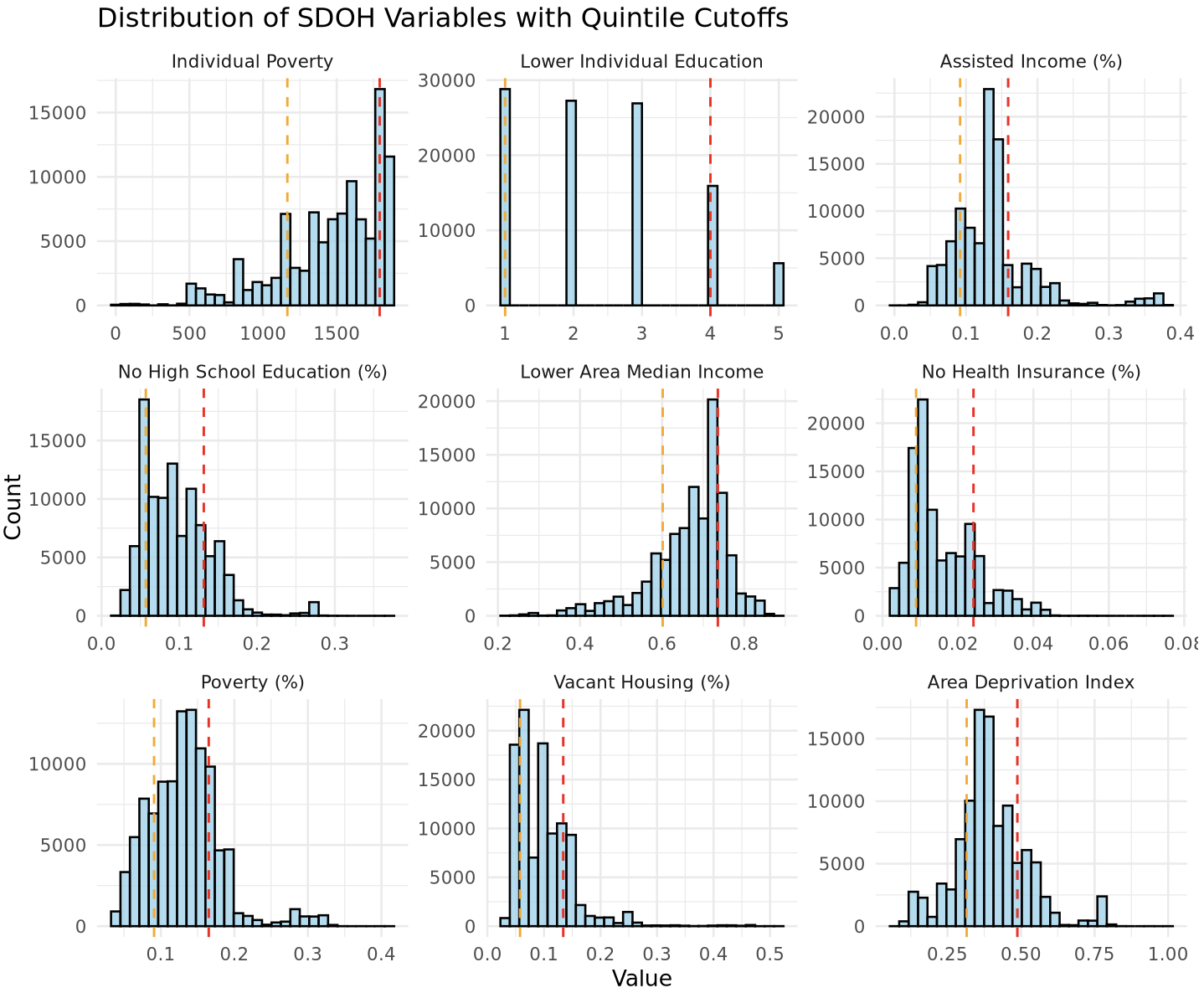

**Supplementary Figure 6:** Distributions of SDoH variables with quantile cutoffs. Distributions of SDoH variables in the direction of increasing social adversity after being z-score normalized. The orange and red dashed lines indicate the cutoffs for Q1 and Q5, respectively, of social adversity. Because cutoffs are made at specific quantile values, there are a different number of samples in each quantile (20905 to 28807 per quantile).

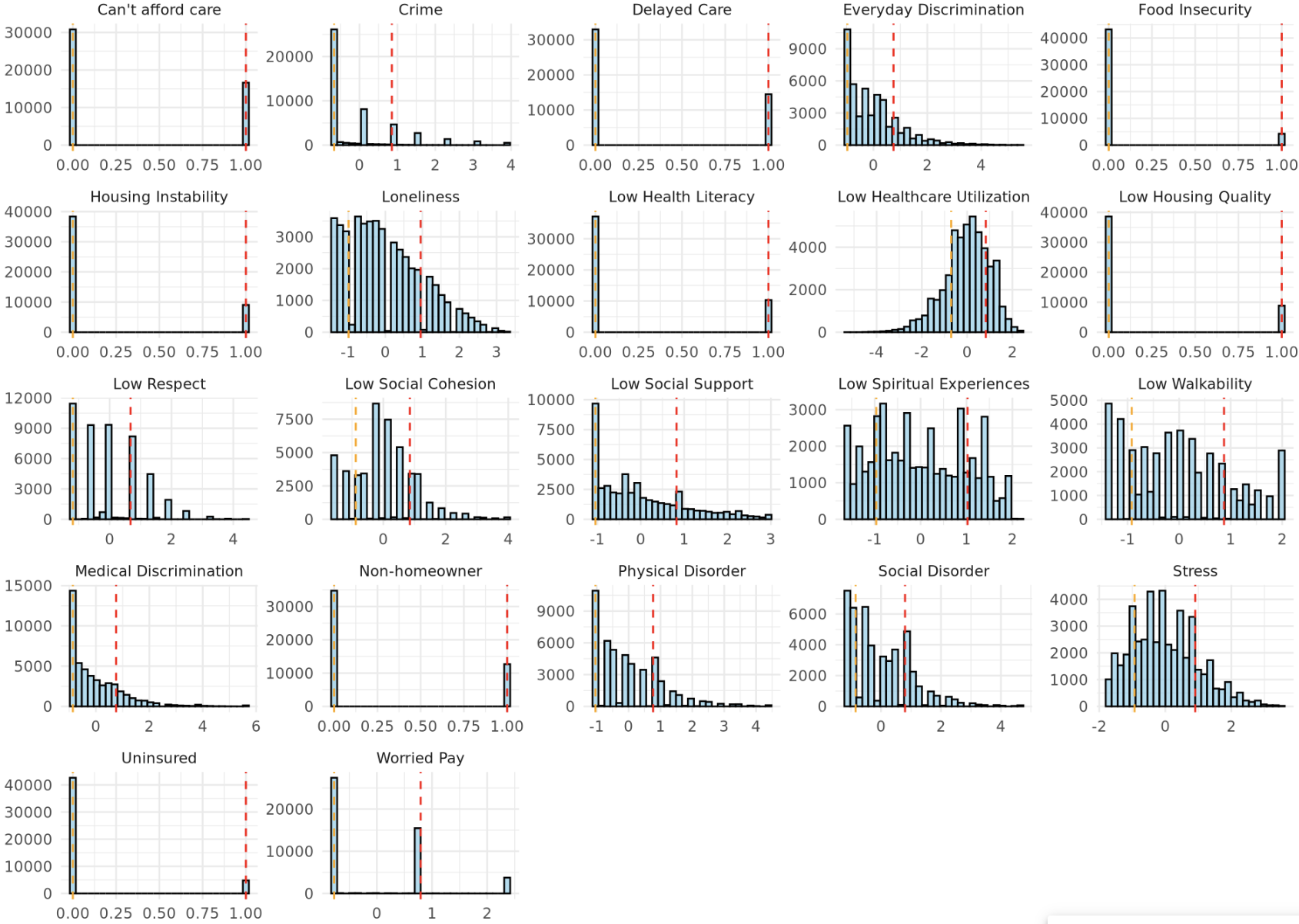

**Supplementary Figure 7:** Distributions of standardized SDoH variables with high/low cutoffs. Distributions of SDoH variables in the direction of increasing social adversity after continuous variables were z-score normalized. The orange and red dashed lines indicate the cutoffs for Q1 and Q5, respectively, of social adversity for continuous traits or binary groups for binary traits. Because cutoffs are made at specific quantile values, there are a different number of samples in each quantile.

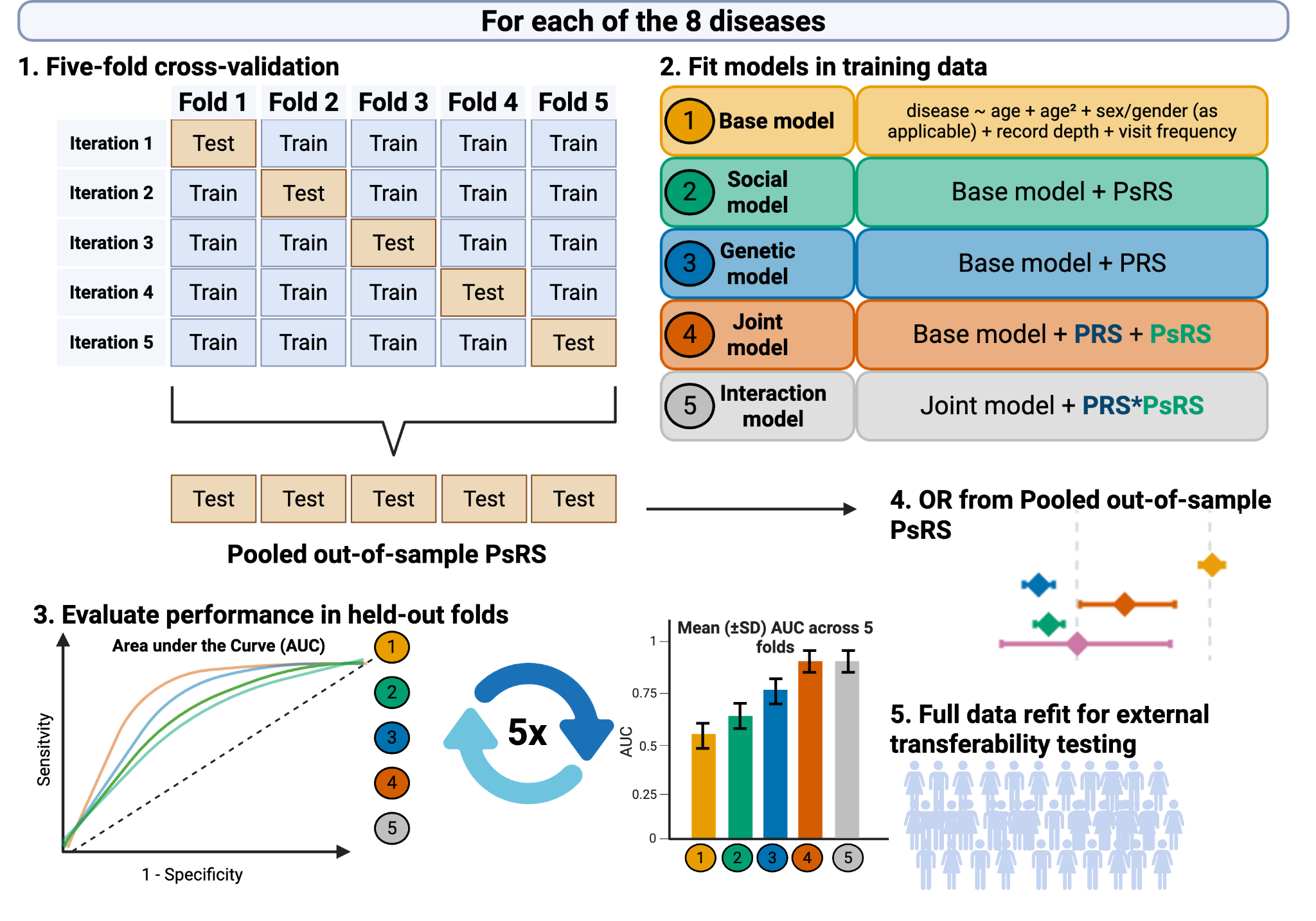

**Supplementary Figure 8:** Graphical depiction of PsRS workflow from randomly partitioning the cohort into 5 folds for 5-fold cross-validation, to fitting models within the training data, using held-out testing data to evaluate models, using pooled out-of-sample PsRS to estimate PsRS OR, to performing a full data refit to obtain model weights for external transferability testing.

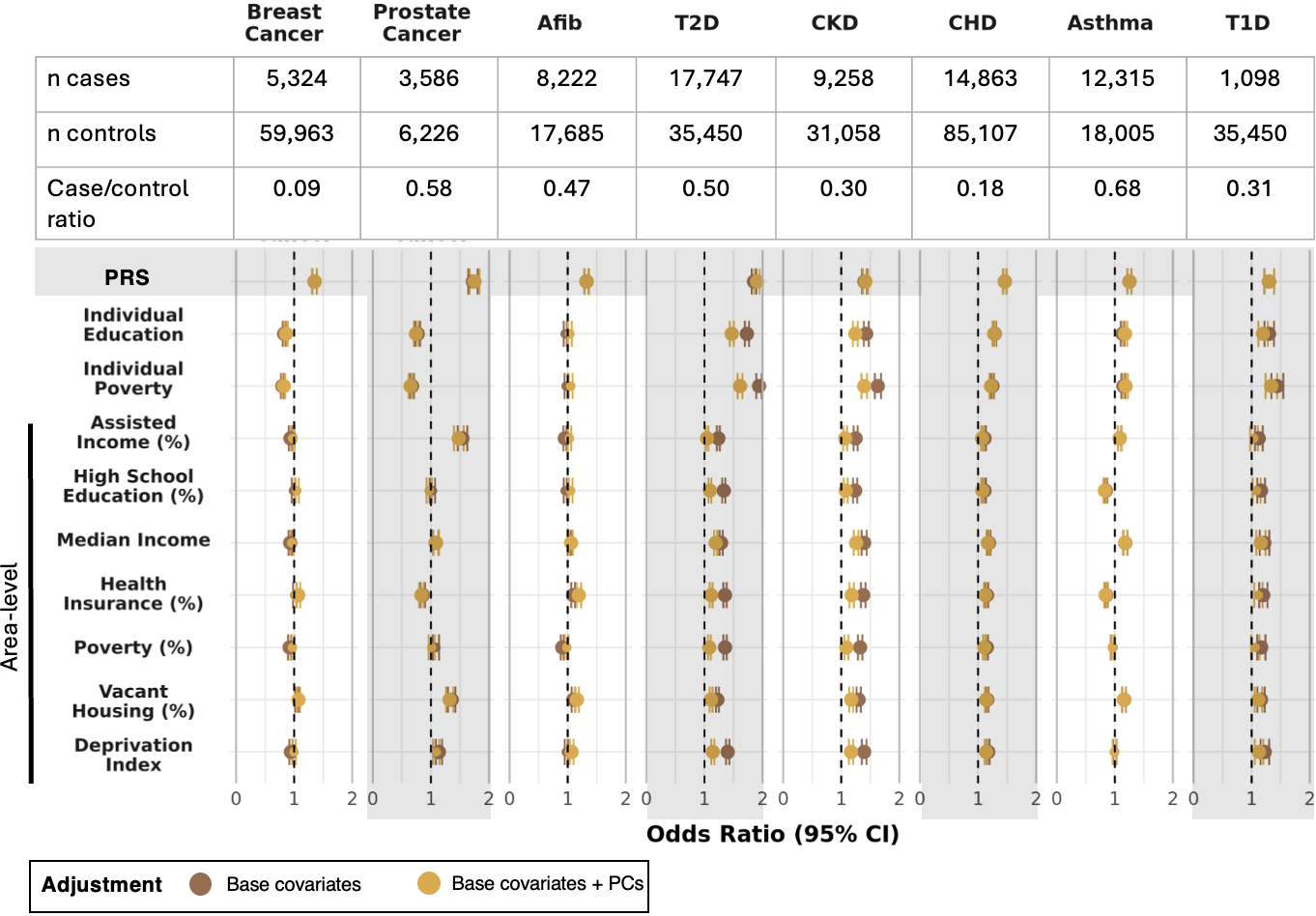

**Supplementary Figure 9:** Forest plots of odds ratios of PRS and SDoH with and without additional covariate adjustment for 16 genetic PCs. For each disease (columns) and SDoH variable (rows), the forest plots display the odds ratio (OR) and 95% confidence interval (CI) for PRS (top row; grey highlight) and SDoH variables with (yellow) and without (brown) adjustment for 16 genetic PCs. A null OR of 1 is indicated with a black vertical dashed line. Point size indicates significance, with larger points indicating statistical significance after Bonferroni correction (P < 6.25 × 10⁻⁴; 0.05/80 comparisons). Case sizes (in thousands) are displayed below each disease and the case/control ratio is displayed below that.

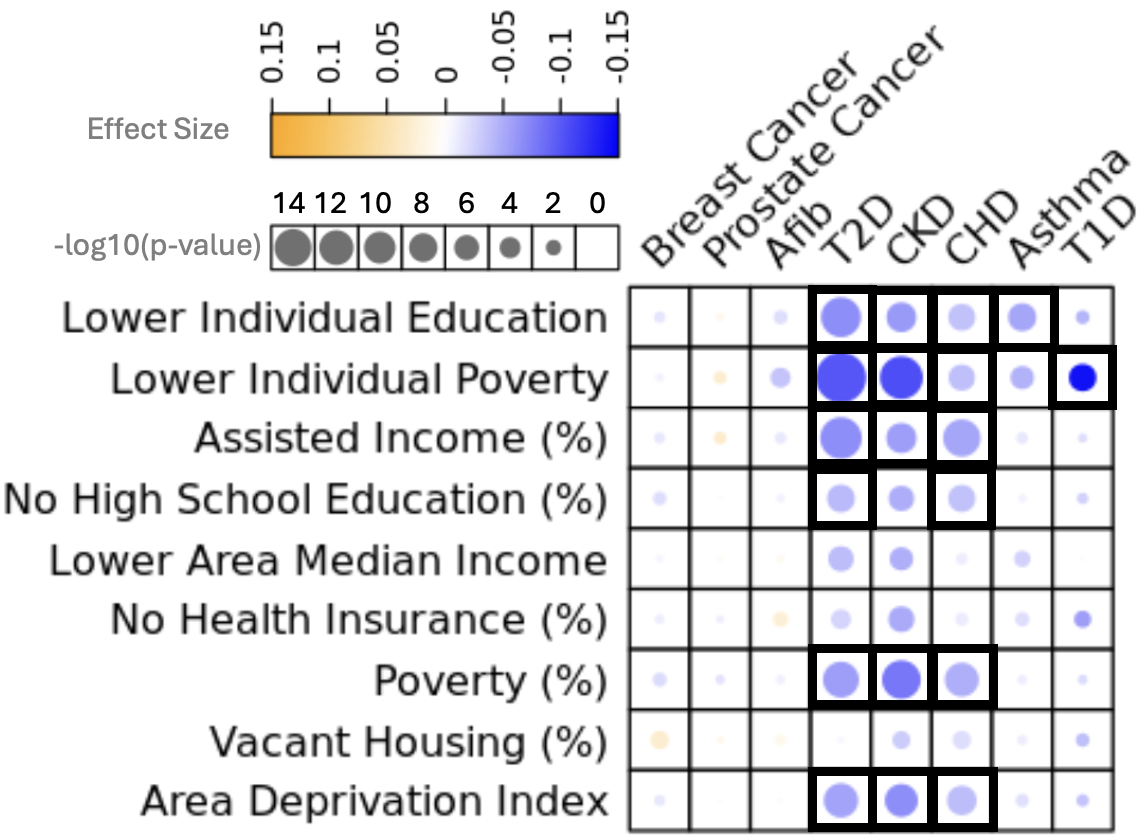

**Supplementary Figure 10:** Bubble heatmap of interactions between SDoH and PRS on disease outcomes. Effect size is displayed via a color gradient, where blue indicates negative interactions. Negative interactions with lower social vulnerability and disease PRS indicate that the association between PRS and disease risk decreases as social vulnerability increases and is stronger at lower levels of social vulnerability. Bubble size denotes p-value significance. Thicker line width indicates interaction terms are significant at a Bonferroni p-value of 6.94e-04 after correcting for 72 SDoH-disease pairs.

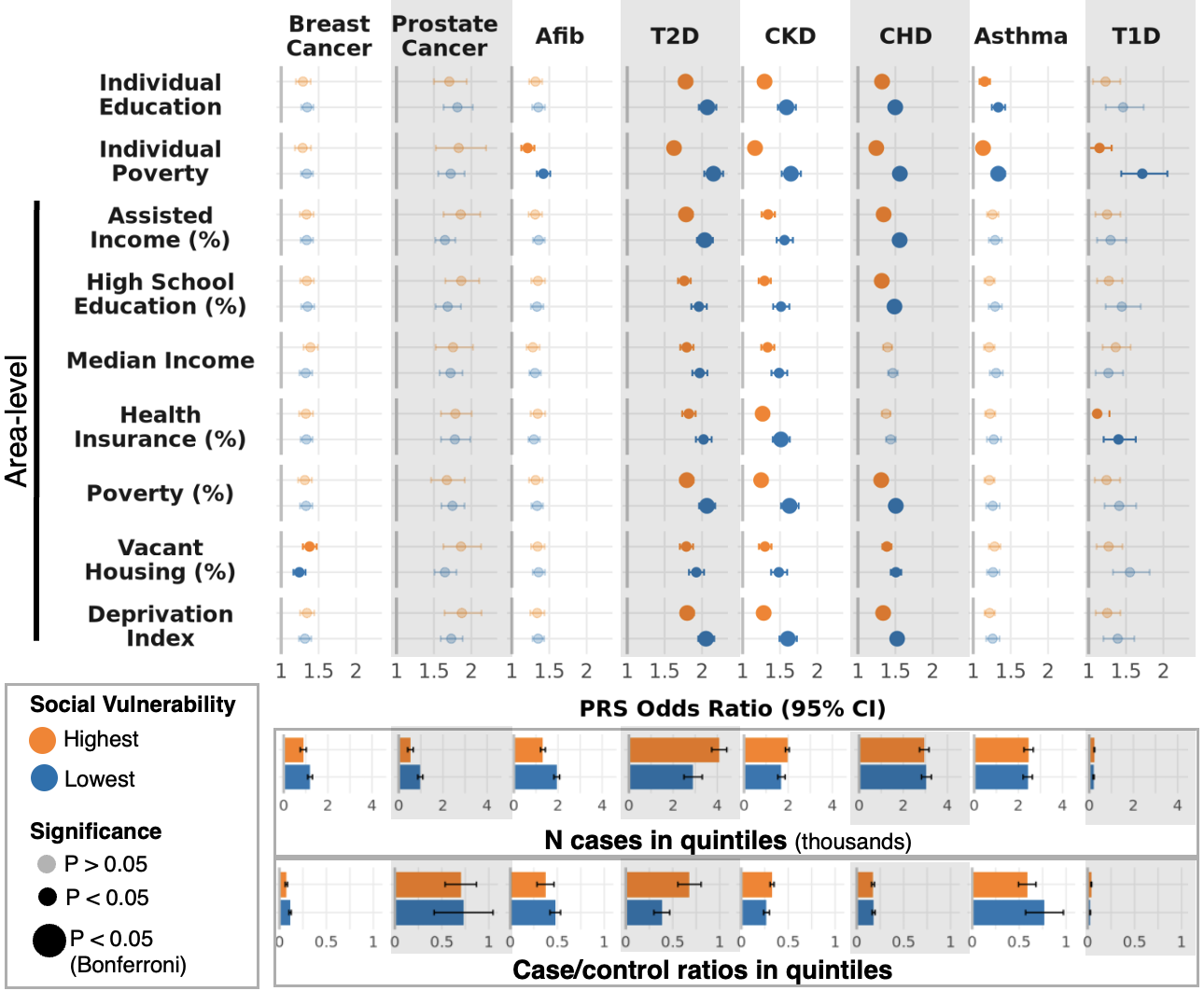

**Supplementary Figure 11:** Forest plots of odds ratios of PRS in SDoH stratified analyses (as in Figure 2), with additional covariate adjustment for 16 genetic PCs. For each disease (columns) and SDoH variable (rows), the forest plots display the odds ratio (OR) and 95% confidence interval (CI) for the PRS in the highest (Q5; orange) and lowest (Q1; blue) social adversity quintiles. Mean case sample sizes and case/control ratios for Q1 and Q5 across SDoH strata are displayed below each disease, with error bars representing ±1 standard deviation across SDoH strata. Case and control sample sizes for each individual disease–SDoH stratum are provided in Supplementary Table 6. Two-sided z-tests were used to assess differences between PRS ORs in the highest and lowest social adversity groups. ORs without significant differences between groups are displayed with increased transparency, whereas comparisons that remained statistically significant after Bonferroni correction (P < 6.94 × 10⁻⁴; 0.05/72 comparisons) are indicated by larger points.

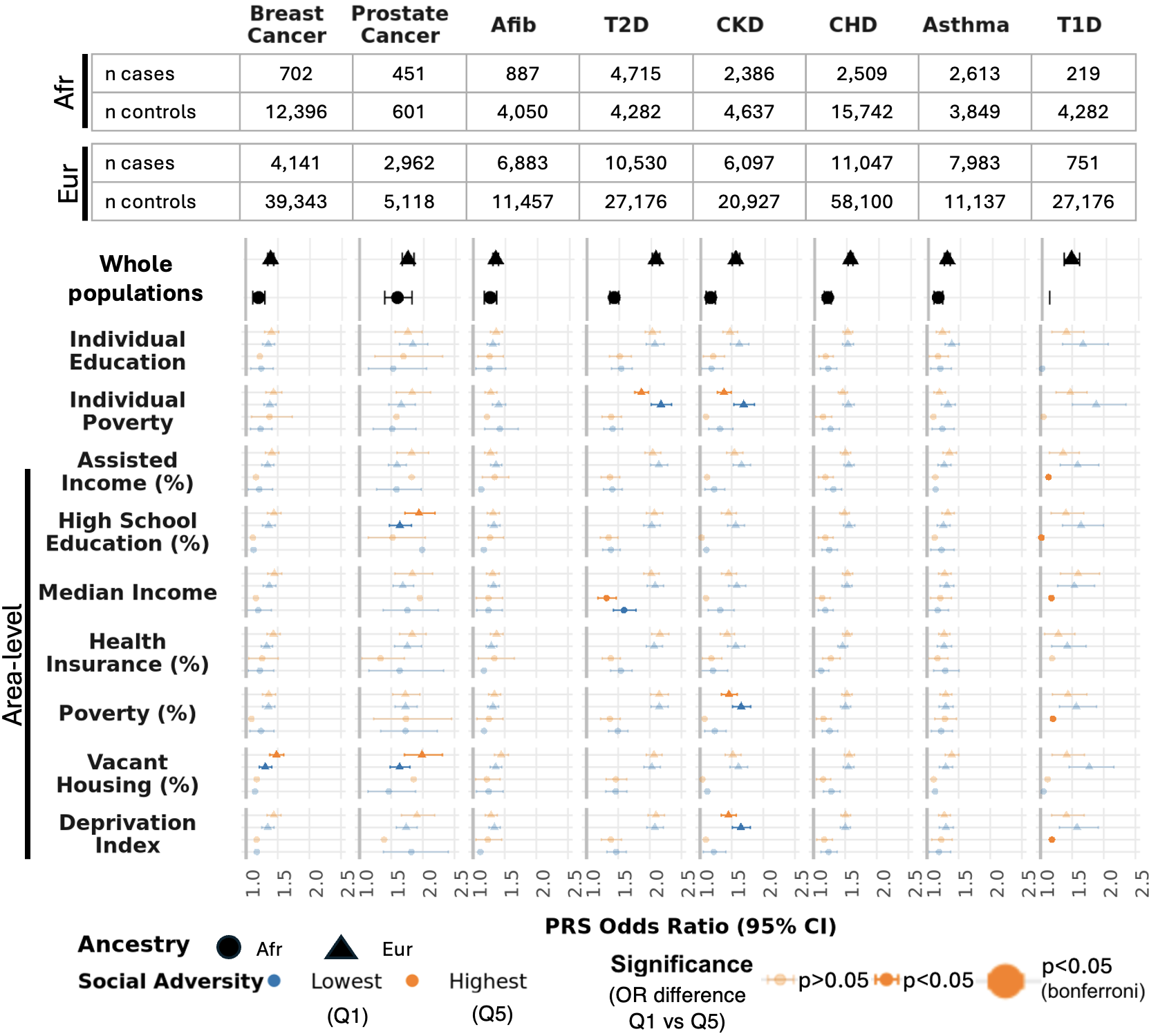

**Supplementary Figure 12:** Forest plots of odds ratios of PRS in SDoH- and continental genetic ancestry-stratified analyses. For each disease (x-axis) and SDoH (y-axis) pair, the plot displays the odds ratio for the PRS in the top and bottom quintile of social adversity within each of the two largest ancestry clusters. The odds ratio of the PRS in the highest adversity group is shown in orange while the odds ratio in the lowest social adversity group is in blue. Analyses conducted in the African-like genetic similarity (Afr) cluster have their odds ratios represented as a dot while those conducted in the European-like genetic similarity (Eur) cluster as a triangle. Error bars represent 95% confidence intervals. A two-sided z-test was used to assess differences between PRS odds ratios in the highest and lowest social adversity quintiles. Odds ratios with non-significant differences are displayed with increased transparency, whereas those that remain statistically significant after Bonferroni correction (p < 6.94 × 10⁻⁴; 0.05/72) are shown with larger point sizes.

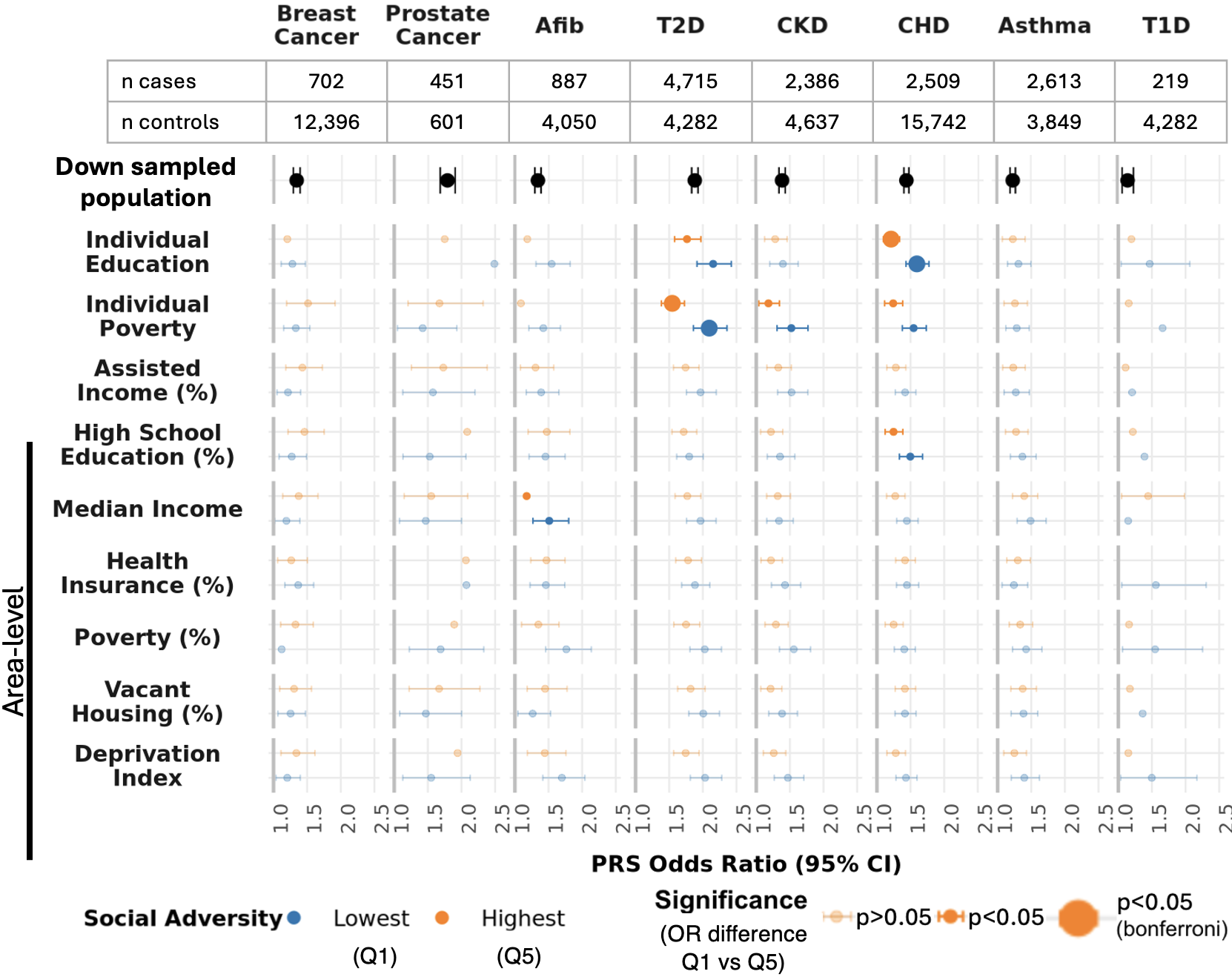

**Supplementary Figure 13:** Forest plots of odds ratios of PRS in SDoH in a randomly downsampled whole population. For each disease (x-axis) and SDoH (y-axis) pair, the plot displays the odds ratio for the PRS in the top and bottom quintile of social adversity within the whole population randomly downsampled to match the size of the smaller Afr-like cluster. The odds ratio of the PRS in the highest adversity group is shown in orange while the odds ratio in the lowest social adversity group is in blue. Error bars represent 95% confidence intervals. A two-sided z-test was used to assess differences between PRS odds ratios in the highest and lowest social adversity quintiles. Odds ratios with non-significant differences are displayed with increased transparency, whereas those that remain statistically significant after Bonferroni correction (p < 6.94 × 10⁻⁴; 0.05/72) are shown with larger point sizes.

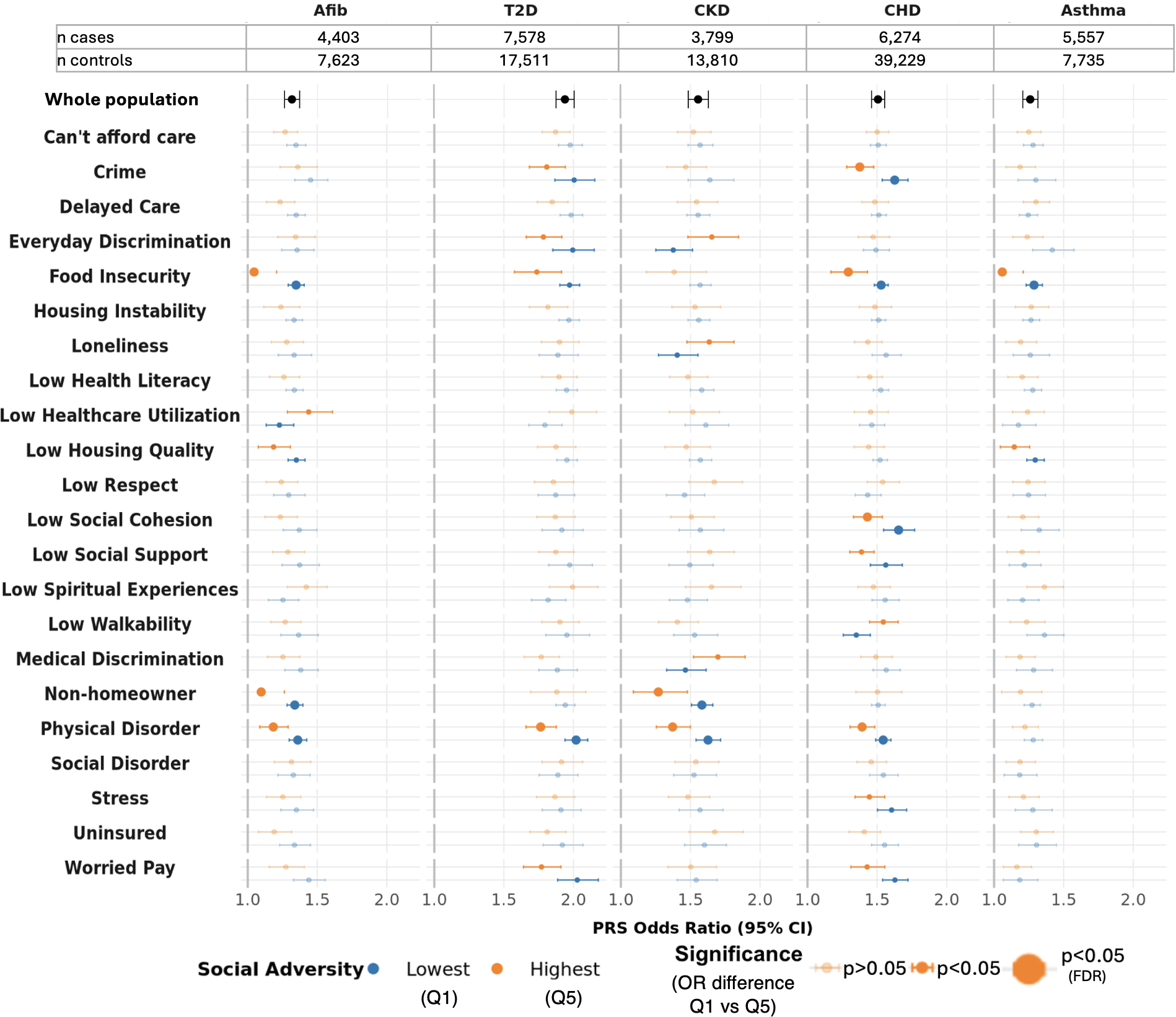

**Supplementary Figure 14:** Forest plots of odds ratios of PRS in SDoH stratified analyses using expanded individual-level SDoH data**.** For each disease (x-axis) and SDoH (y-axis) pair, the plot displays the OR for the PRS in the top and bottom quintile of social vulnerability after controlling for poverty and education. Above these stratified results are the PRS ORs and the case-control sample sizes in the whole cohort. For the SDoH-stratified analyses, the OR of the PRS in the highest adversity group is shown in orange while the OR in the lowest social vulnerability group is in blue. Case-control sizes within these quantiles are available in Supplementary Table 8. Error bars represent 95% confidence intervals. A two-sided z-test was used to assess differences between PRS ORs in the highest and lowest social vulnerability quintiles or groups for binary variables. ORs with non-significant differences are displayed with increased transparency, whereas those that remain statistically significant after FDR correction are shown with larger point sizes.

.

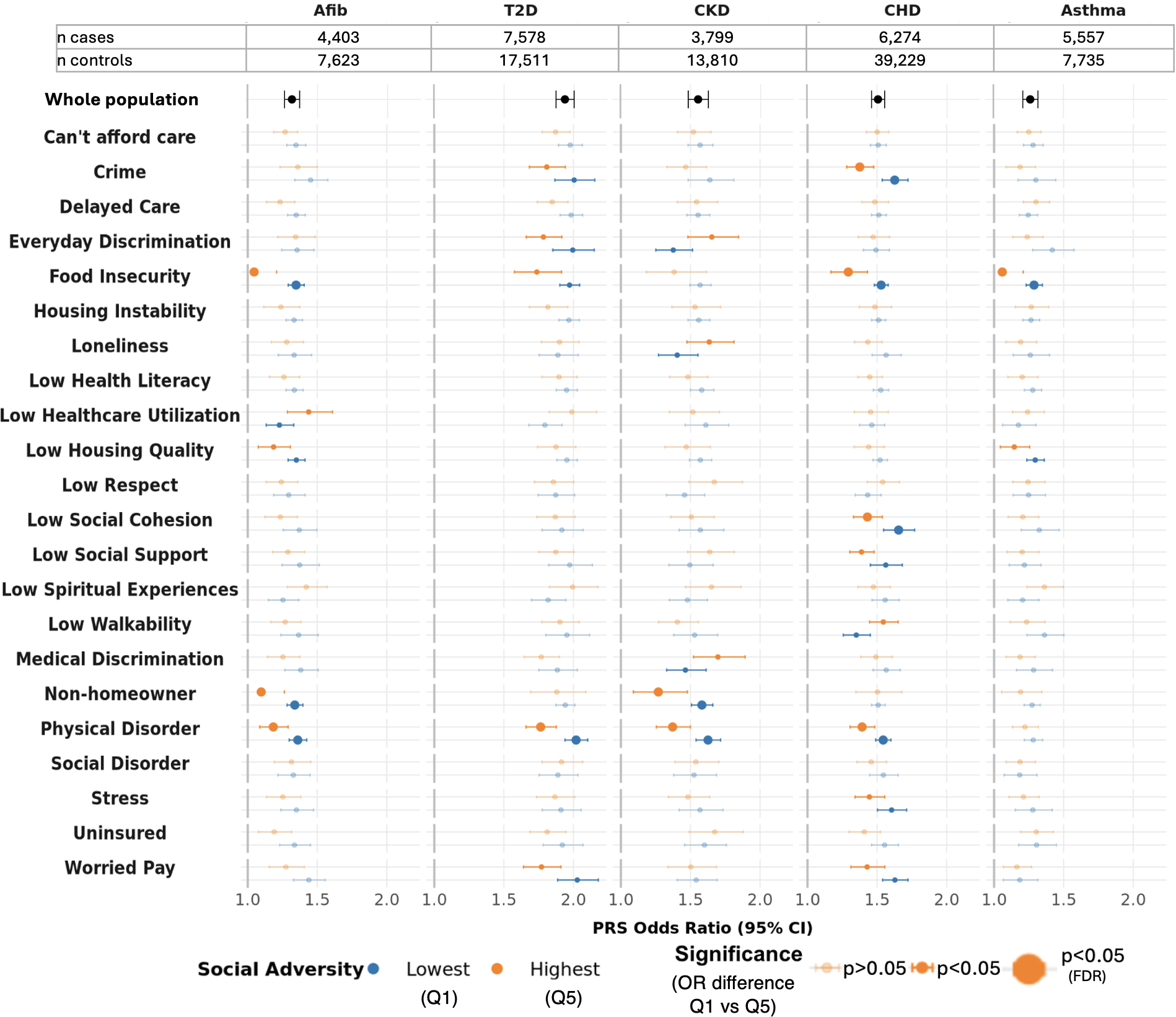

**Supplementary Figure 15:** Forest plots of odds ratios of PRS in SDoH stratified analyses using richer individual-level SDoH data with additional adjustment for genetic PCs as covariates. For each disease (x-axis) and SDoH (y-axis) pair, the plot displays the odds ratio for the PRS in the top and bottom quintile of social adversity after controlling for poverty, education, and 16 genetic PCs. The odds ratio of the PRS in the highest adversity group is shown in orange while the odds ratio in the lowest social adversity group is in blue. Error bars represent 95% confidence intervals. A two-sided z-test was used to assess differences between PRS odds ratios in the highest and lowest social adversity quintiles or groups for binary variables. Odds ratios with non-significant differences are displayed with increased transparency, whereas those that remain statistically significant after FDR correction are shown with larger point sizes.

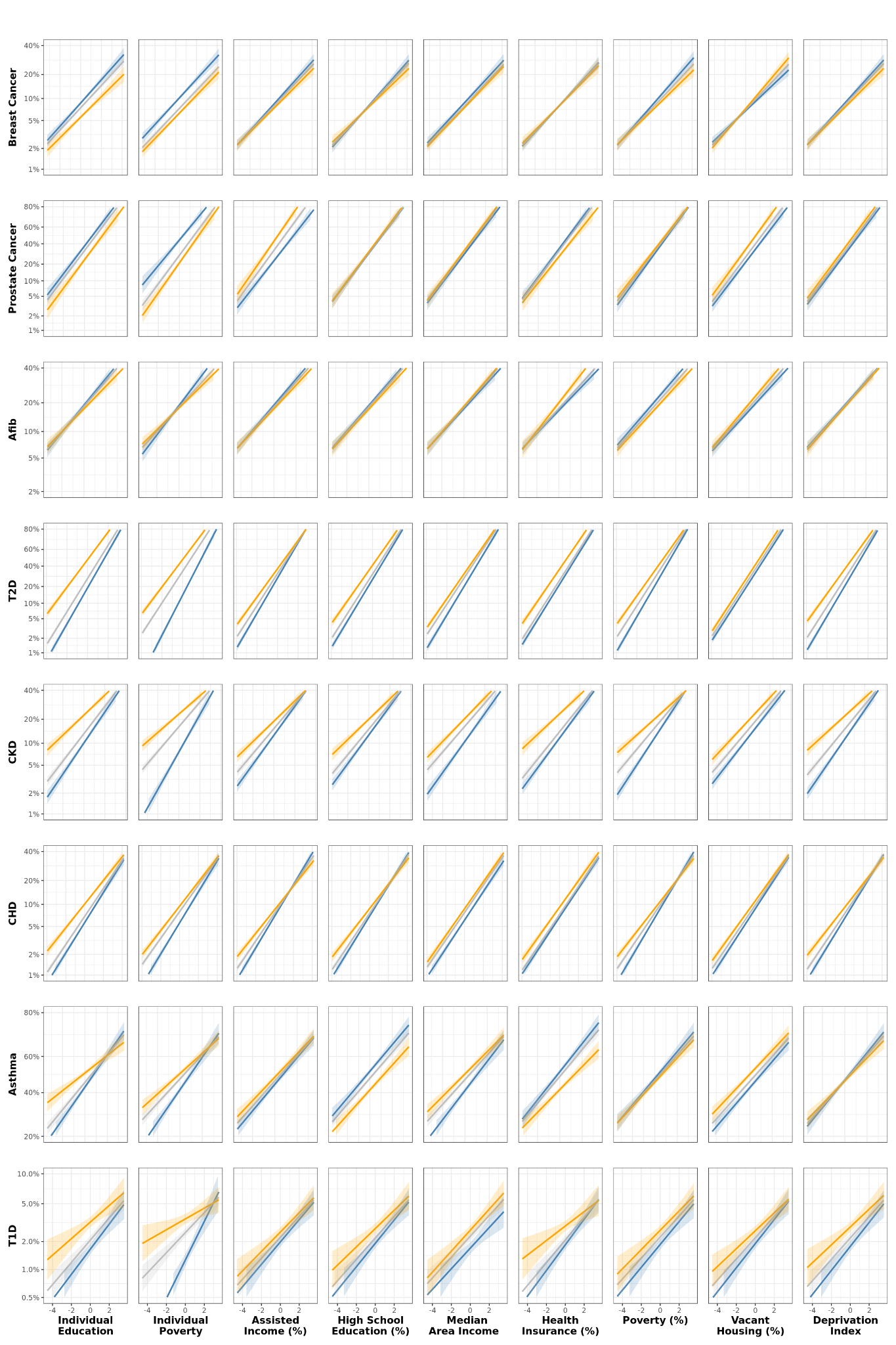

**Supplementary Figure 16:** Grid of conditional PRS effects across SDoH stratum. For each SDoH-disease pair, the PRS increases linearly across the x-axis, while the predicted probability of disease is on a logit scale on the y-axis. Three effect curves represent low (Q1; blue), intermediate (Q2–Q4; grey), and high (Q5; orange) social adversity. Shaded areas denote 95% confidence intervals.

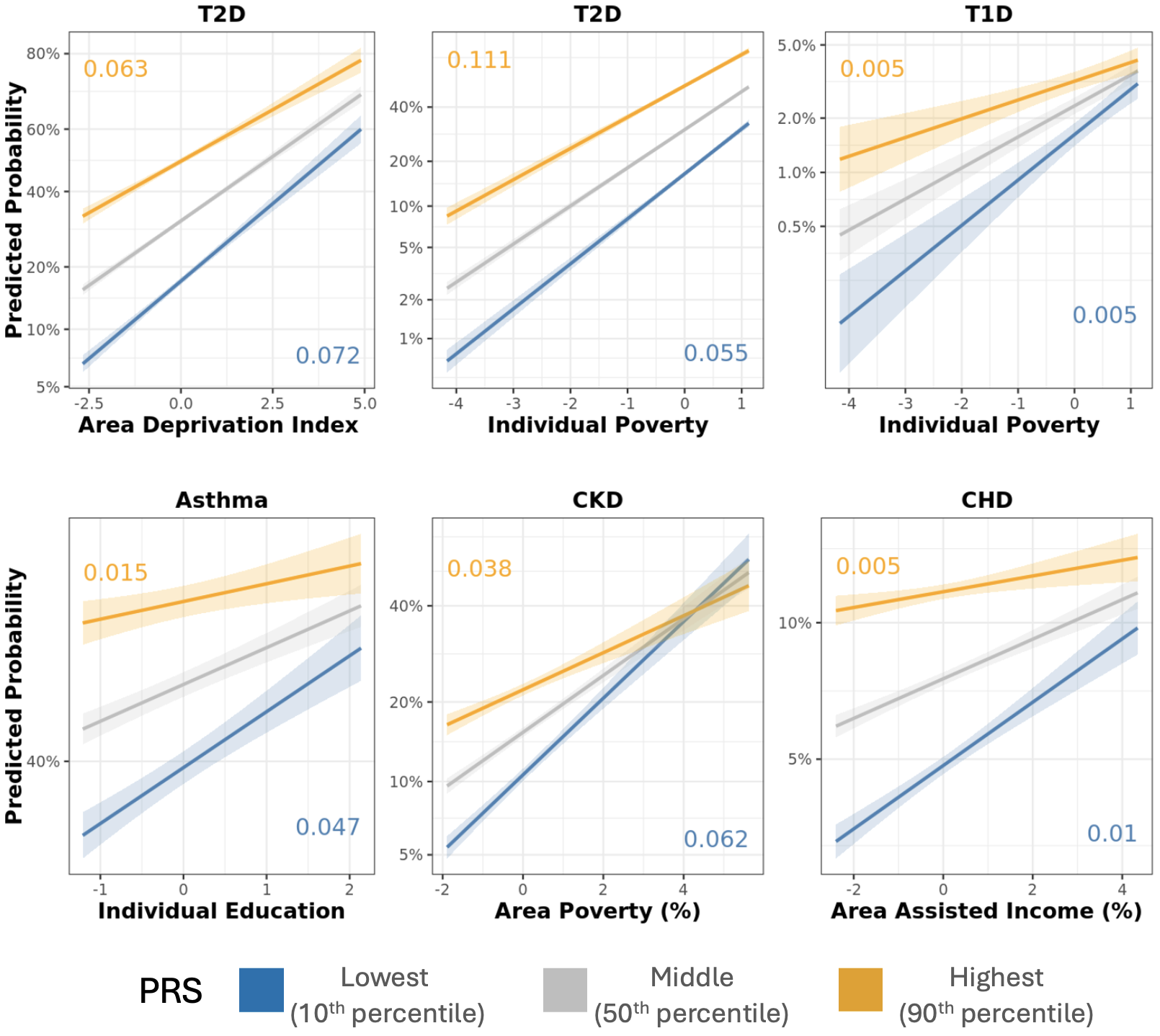

**Supplementary Figure 17:** Effects plots for representative SDoH-disease pairs with significant interactions. For each SDoH-disease pair, the social variable increases linearly across the x-axis, while the predicted probability of disease (based on case-control algorithms in AoU) is on a logit scale on the y-axis. Three effect curves represent low (10^th^ percentile; blue), intermediate (50^th^ percentile; grey), and high (90^th^ percentile; orange) PRS. Shaded areas denote 95% confidence intervals, with their slopes annotated in corresponding colors.

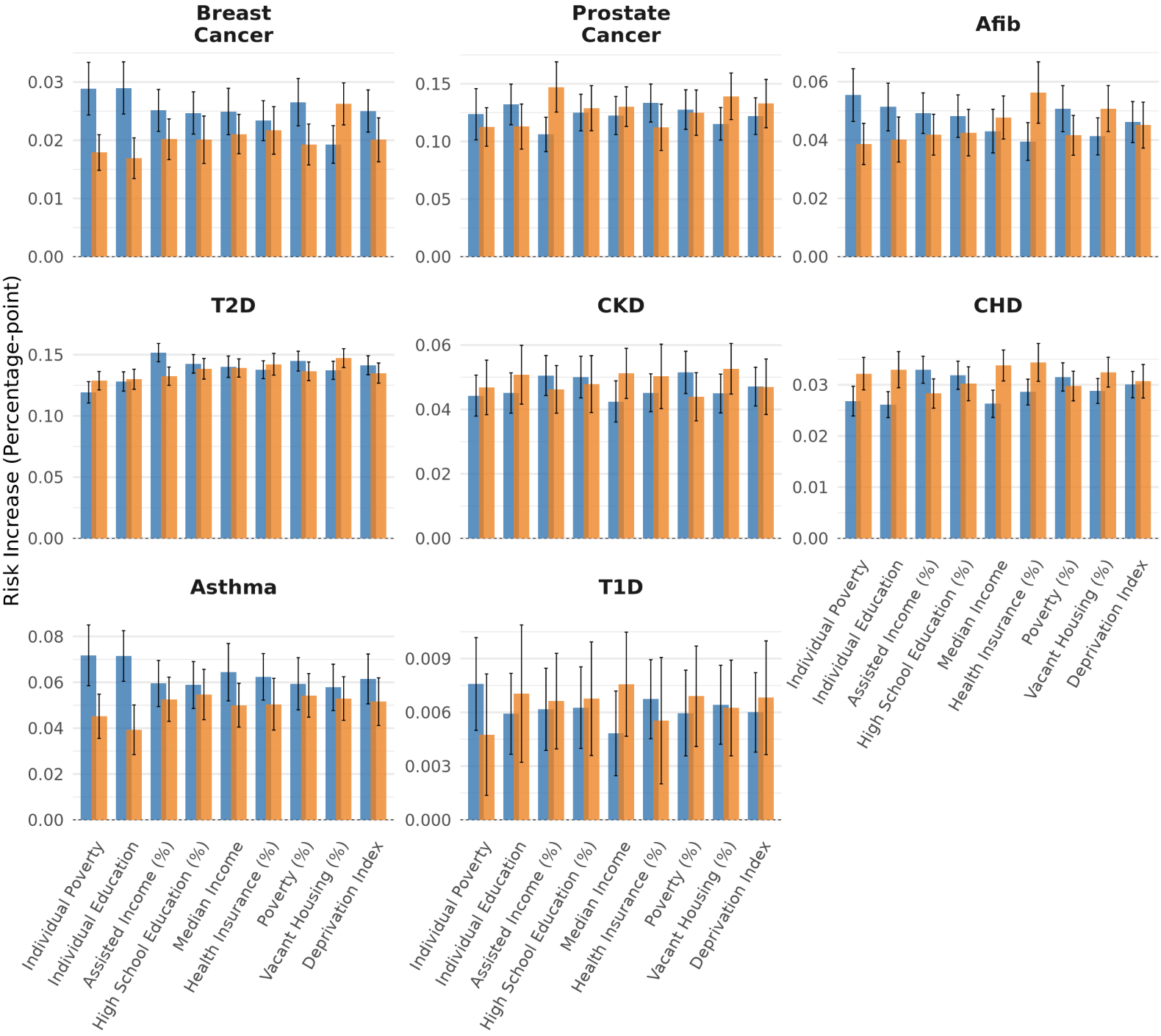

**Supplementary Figure 18:** Relative risk increase associated with PRS for SDoH across eight diseases Change in relative risk of disease per one SD change in PRS for representative individuals in the 10^th^ (blue; lower social risk) and 90^th^ (orange; higher social risk) percentile of social risk across eight diseases. Error bars represent 95% confidence intervals for the relative risk increase.

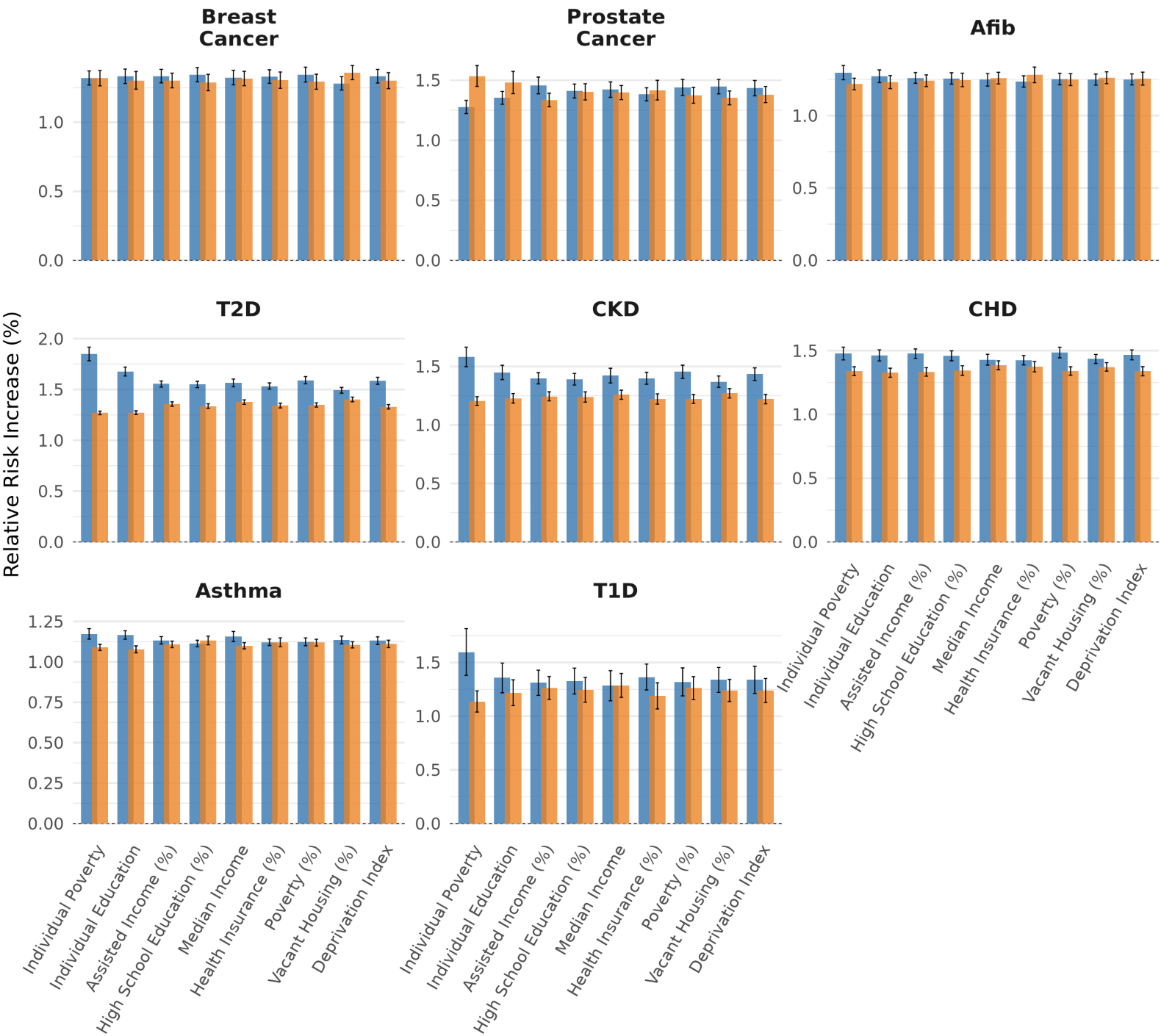

**Supplementary Figure 19:** Model-predicted change in disease probability per 1 SD increase in PRS across levels of social risk. Change in model-predicted disease probability, expressed in percentage points, per 1 SD increase in PRS for representative individuals at the 10th (blue; lower social risk) and 90th (orange; higher social risk) percentiles of social risk across eight diseases. Error bars represent 95% confidence intervals.

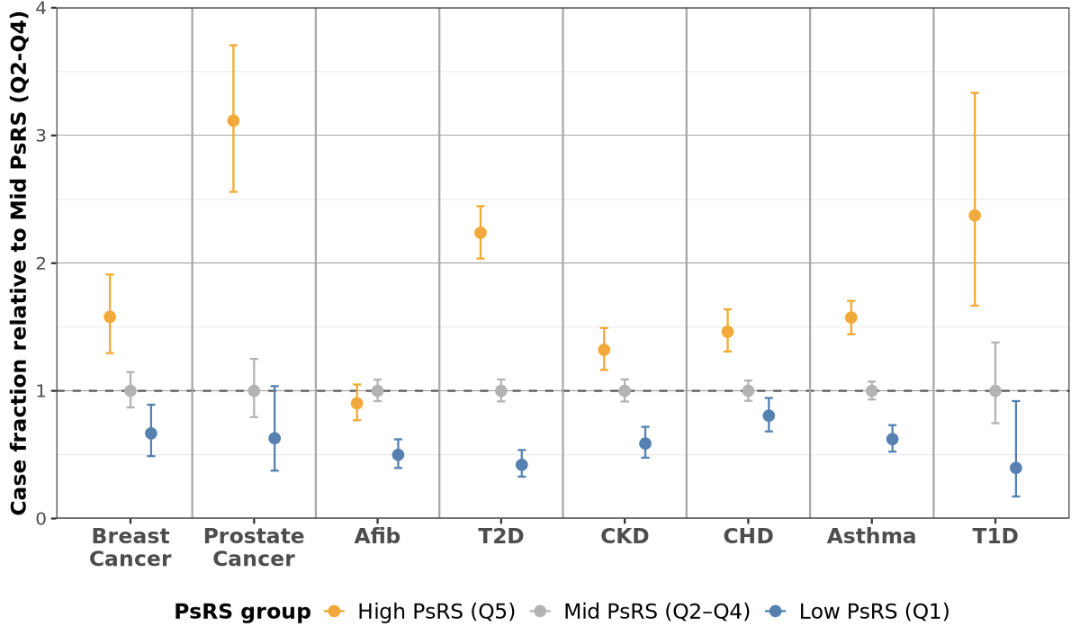

**Supplementary Figure 20:** Dot plot of AoU case fraction among individuals in the lowest genetic risk decile (as opposed to highest in Figure 5), stratified by PsRS quantile. Case fractions are normalized within each disease to the middle PsRS group (Q2-Q4), such that values represent relative case fraction compared with the middle PsRS strata. These estimates reflect AoU case/control phenotype status, which uses stringent control criteria, and should not be interpreted as population disease prevalence. Error bars represent the 95% Wilson confidence interval for the observed case proportion, accounting for binomial sampling uncertainty. Blue dots represent low (Q1) PsRS, grey mid (Q2-4) PSRS, and orange represent high (Q5) social vulnerability.

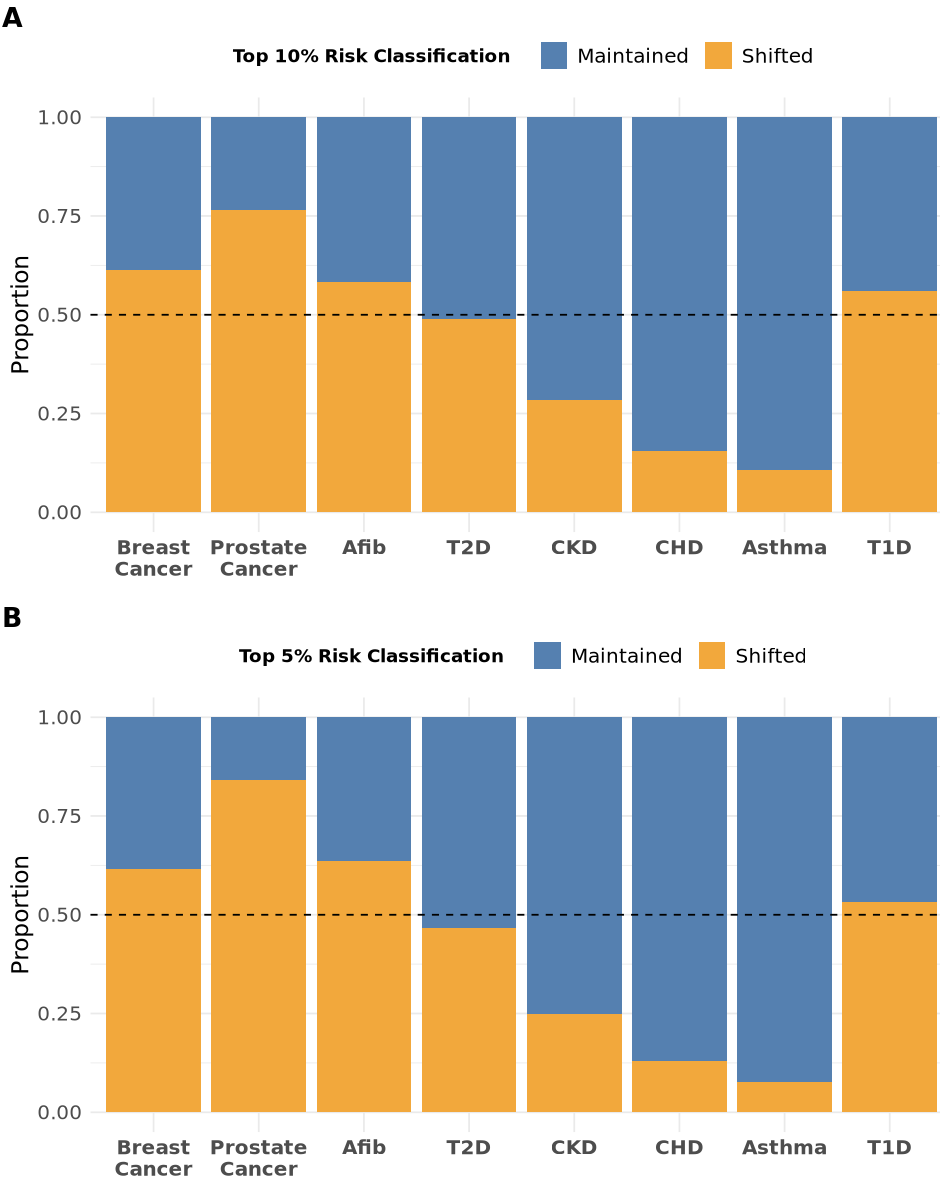

**Supplementary Figure 21:** Top risk classification proportion maintained vs shifted

100% stacked bar charts indicating the proportion of individuals who move in or out of the top risk decile (shifted) vs remain (maintained) when PsRS is added to genetic risk estimates (age, age², sex/gender, record depth, and visit frequency, PRS). The grey dotted indicates that 50% of the top risk classification was the same population and 50% was different after additionally considering PsRS. **A)** Top risk group is classified as top 10% and **B)** top risk group is classified as top 5%. These illustrative high-risk threshold were defined within the analytic cohort and therefore does not represent the top 10% or 5% of risk in the underlying population. The proportion of individuals that changed risk classification ranged from 6% for asthma to 62% for prostate cancer (mean 31% ± 20%); results were similar using the top 5% threshold (mean 32% ± 23%).

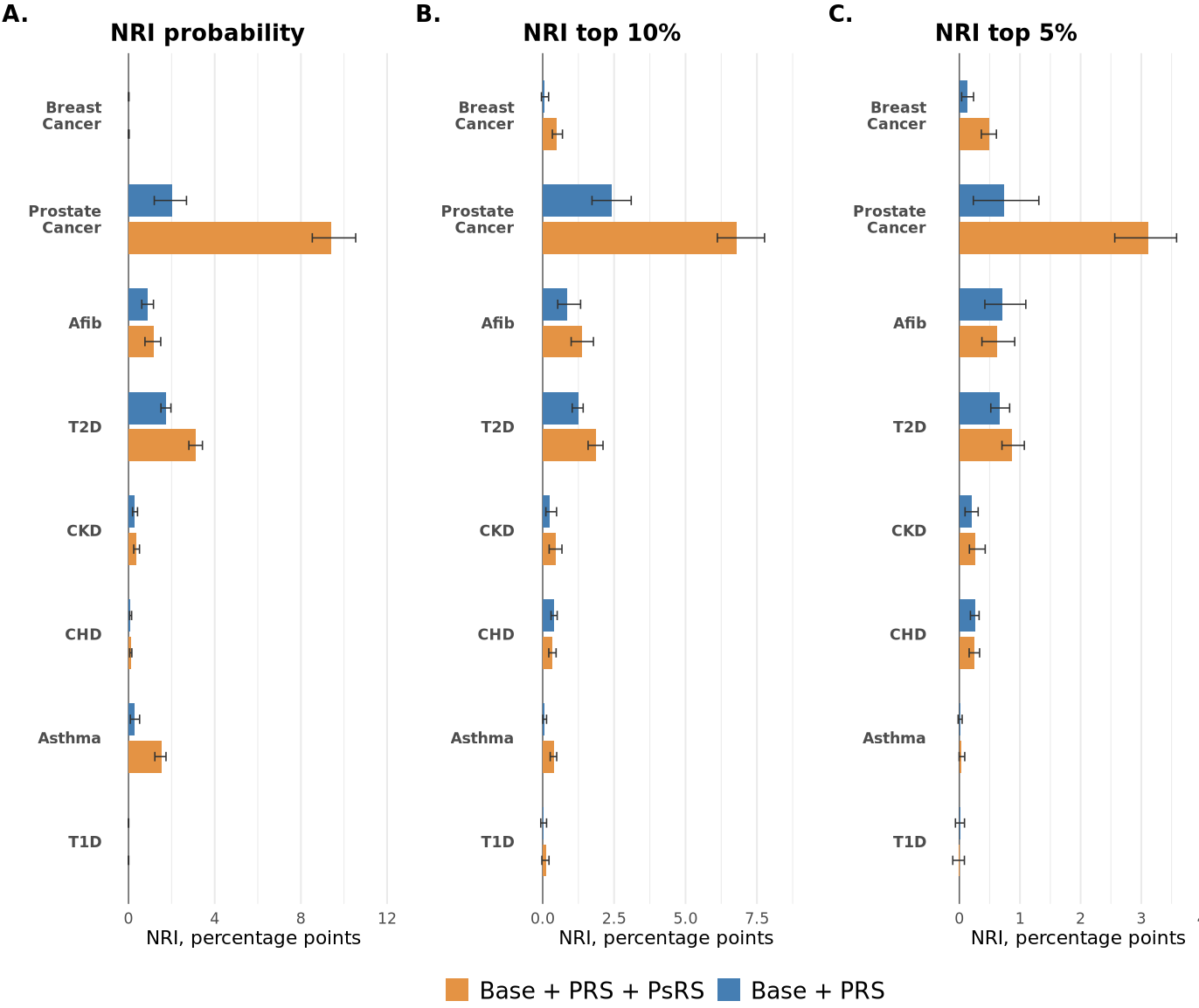

**Supplementary Figure 22:** Comparison of net reclassification improvement (NRI) between models including PRS and those additionally incorporating PsRS relative to the Base model. Positive NRI values indicate net improvement in reclassification by the expanded model. **A)** Threshold-free predicted probabilities. **B)** Top 10% predicted-risk categories. **C)** Top 5% predicted-risk categories. Prostate cancer showed substantial improvement across continuous and top 10% and 5% threshold-based NRI, with more modest improvements for T2D and asthma. For prostate cancer, adding PsRS to the Genetic model yielded a continuous NRI of 0.074, corresponding to a 7.4 percentage-point net improvement in risk classification. Using top-10% and top-5% risk thresholds, the corresponding improvements were 4.4 and 2.4 percentage points, respectively.

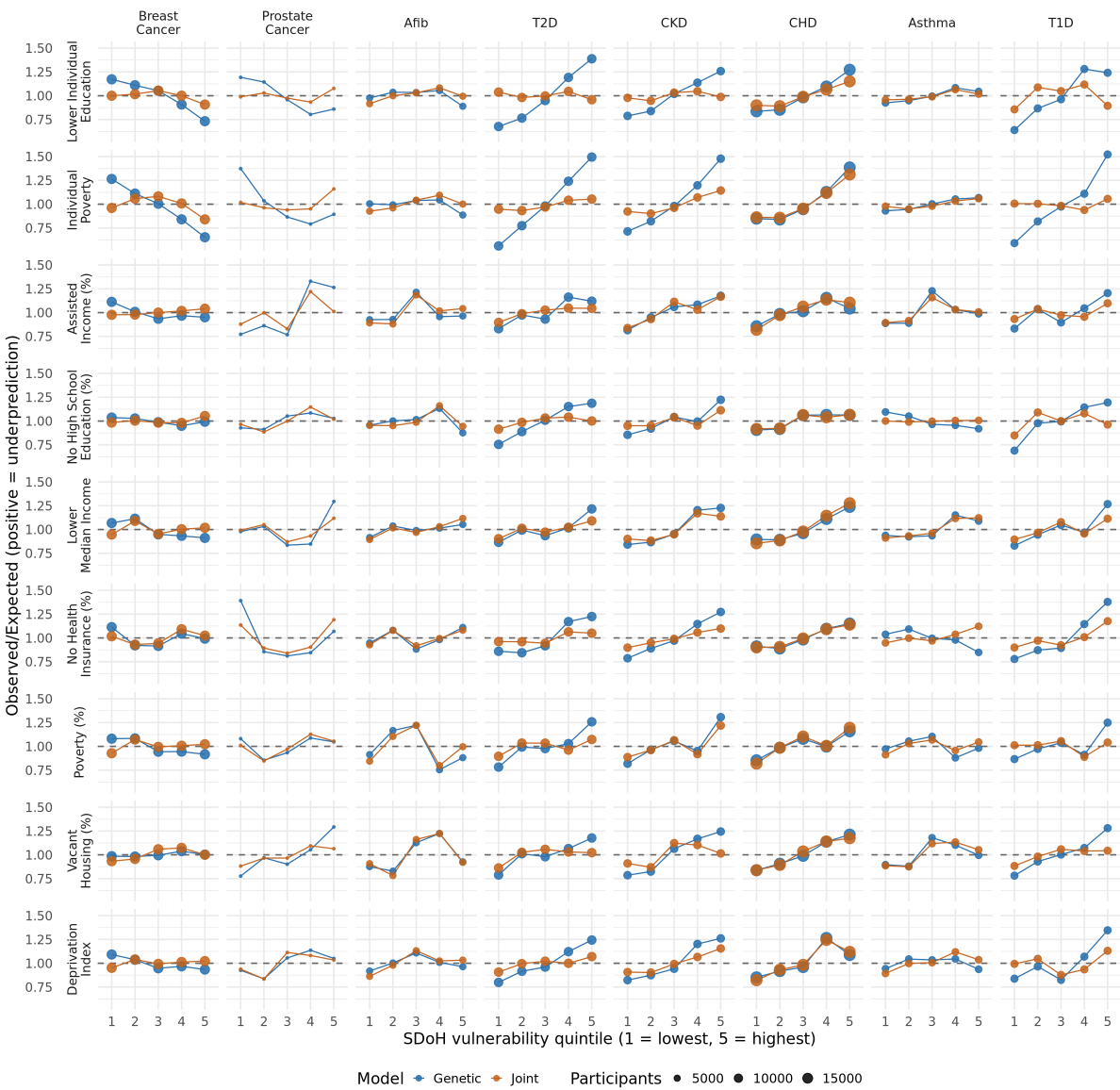

**Supplementary Figure 23:** Observed to expected ratio by SDoH quintile and disease for genetic (blue) and joint (vermillion) models without PC adjustment. An observed/expected ratio of 1 would indicate agreement, values above 1 indicate underprediction, and values below 1 indicate overprediction.

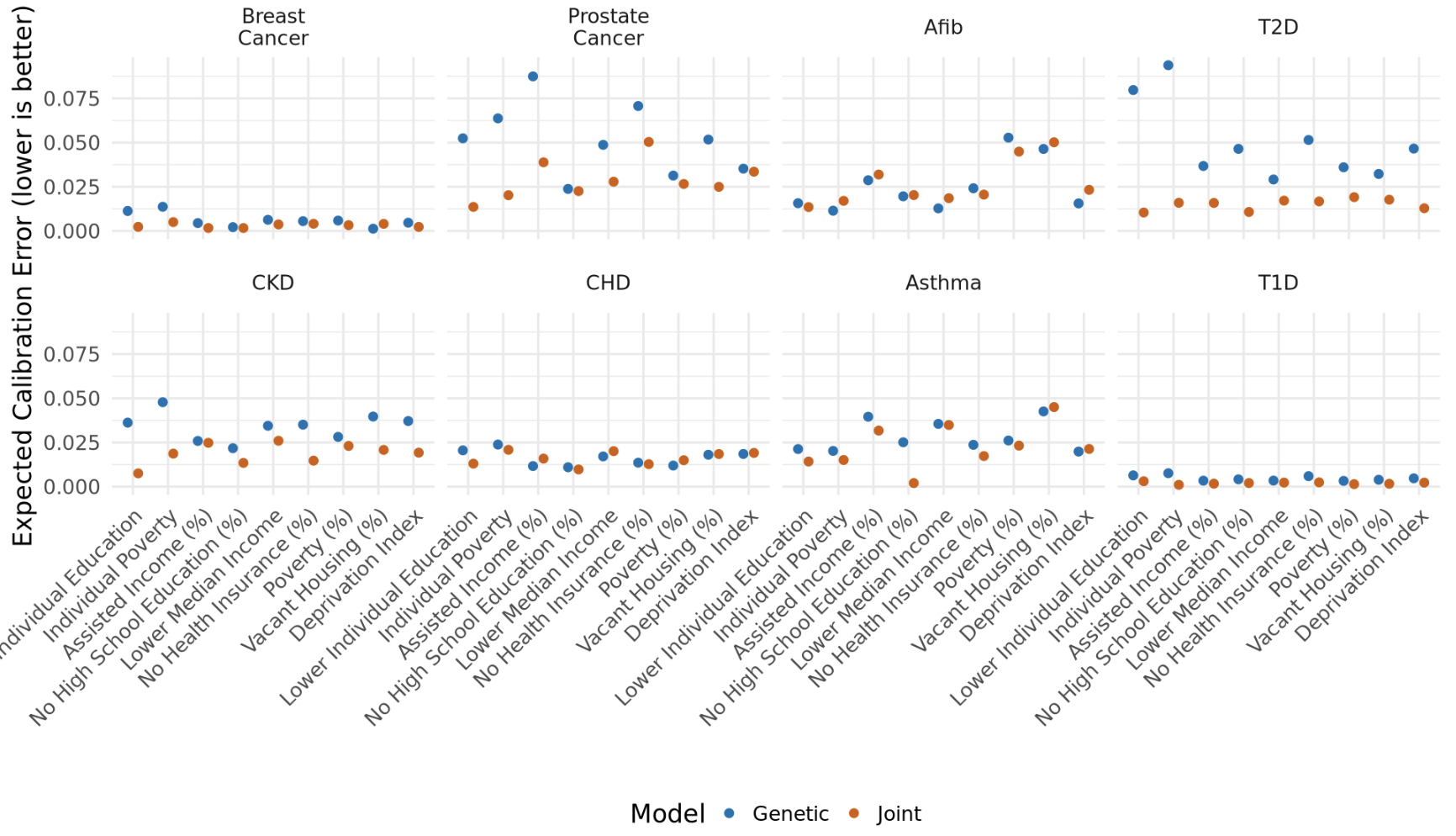

**Supplementary Figure 24:** Expected calibration error (ECE) by disease and SDoH variable for genetic (blue) and joint (vermillion) models without PC adjustment. Lower ECEs are better.

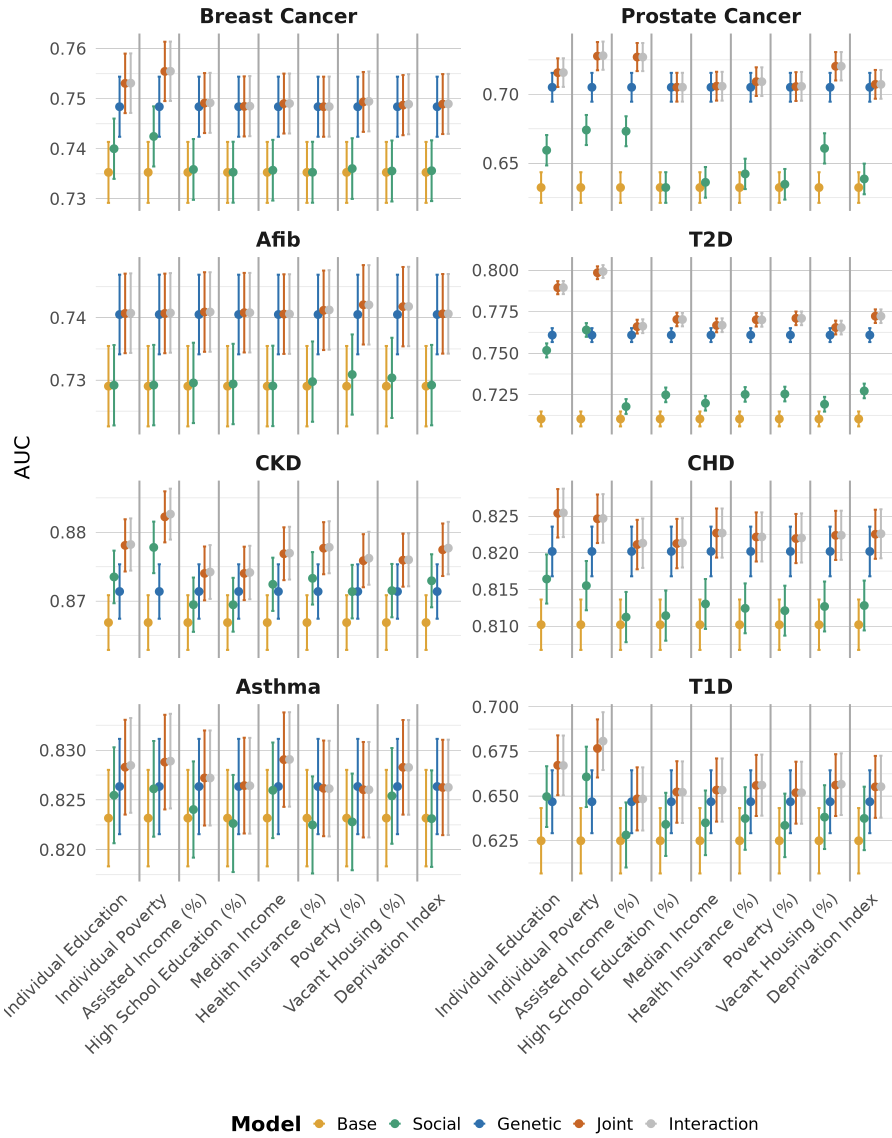

**Supplementary Figure 25:** Comparison of model discrimination (AUC) across diseases for each SDoH measure alone as the social vulnerability metric. The Base model includes demographic and data availability covariates (age, age², sex/gender, record depth, and visit frequency). The Social model extends the Base model by adding corresponding SDoH, while the Genetic model adds PRS. The Joint model includes both PRS and the SDoH, and the Interaction model additionally includes a PRS × SDoH interaction term.

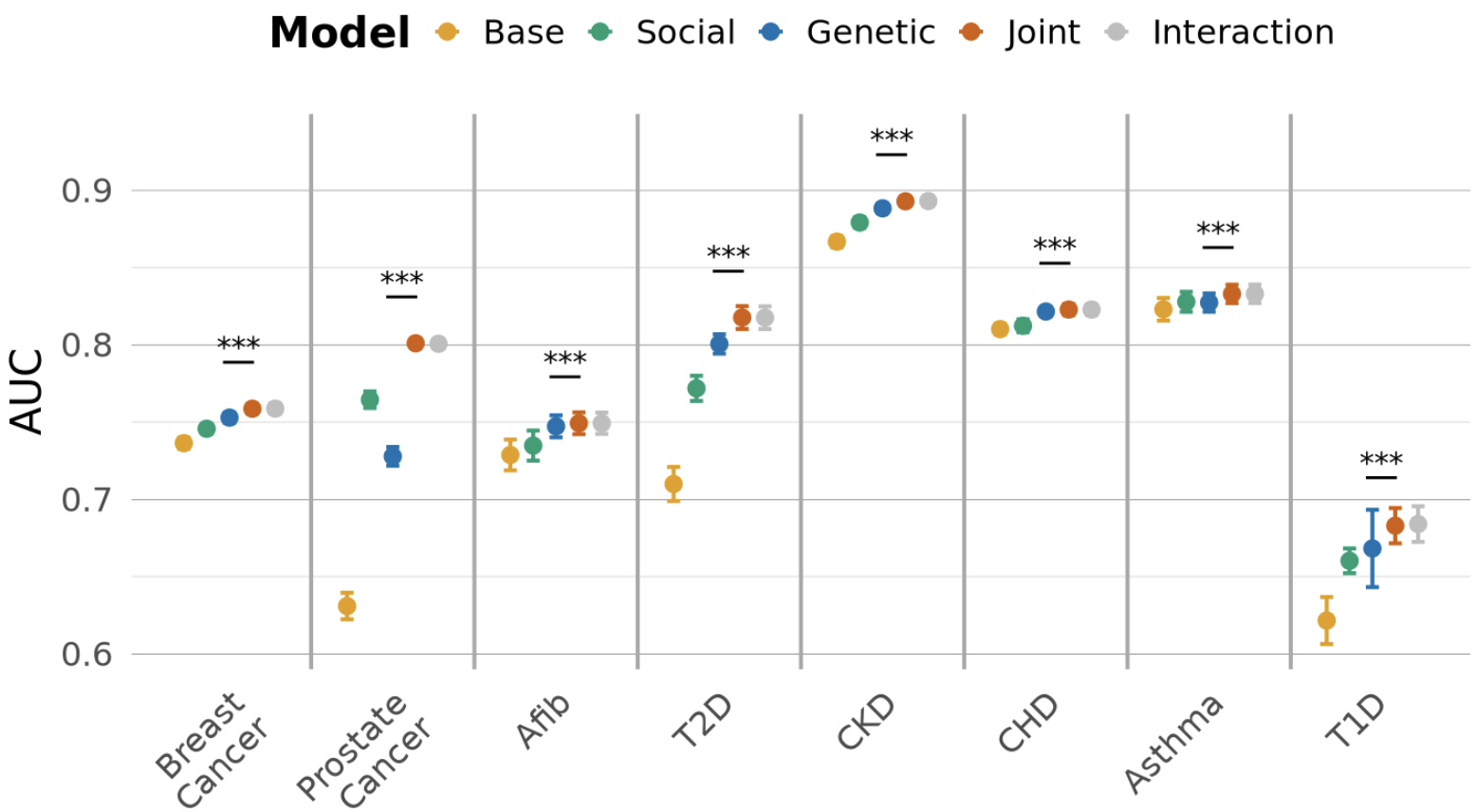

**Supplementary Figure 26:**  Comparison of mean model discrimination (AUC) and SD across five-fold cross validation for eight diseases (as in Figure 6), while additionally controlling for 16 genetic PCs. The Base model includes demographic and data availability covariates (age, age², sex/gender, record depth, and visit frequency). The Social model extends the Base model by adding PsRS, while the Genetic model adds PRS and 16 genetic PCs. The Joint model adds PsRS to the Genetic model, and the Interaction model additionally includes a PRS × PsRS interaction term. Significance comparisons are made between the Genetic and Joint model with 5000 bootstrap resamples. Three stars indicate p < 0.00020 (5000 bootstrap resolution).

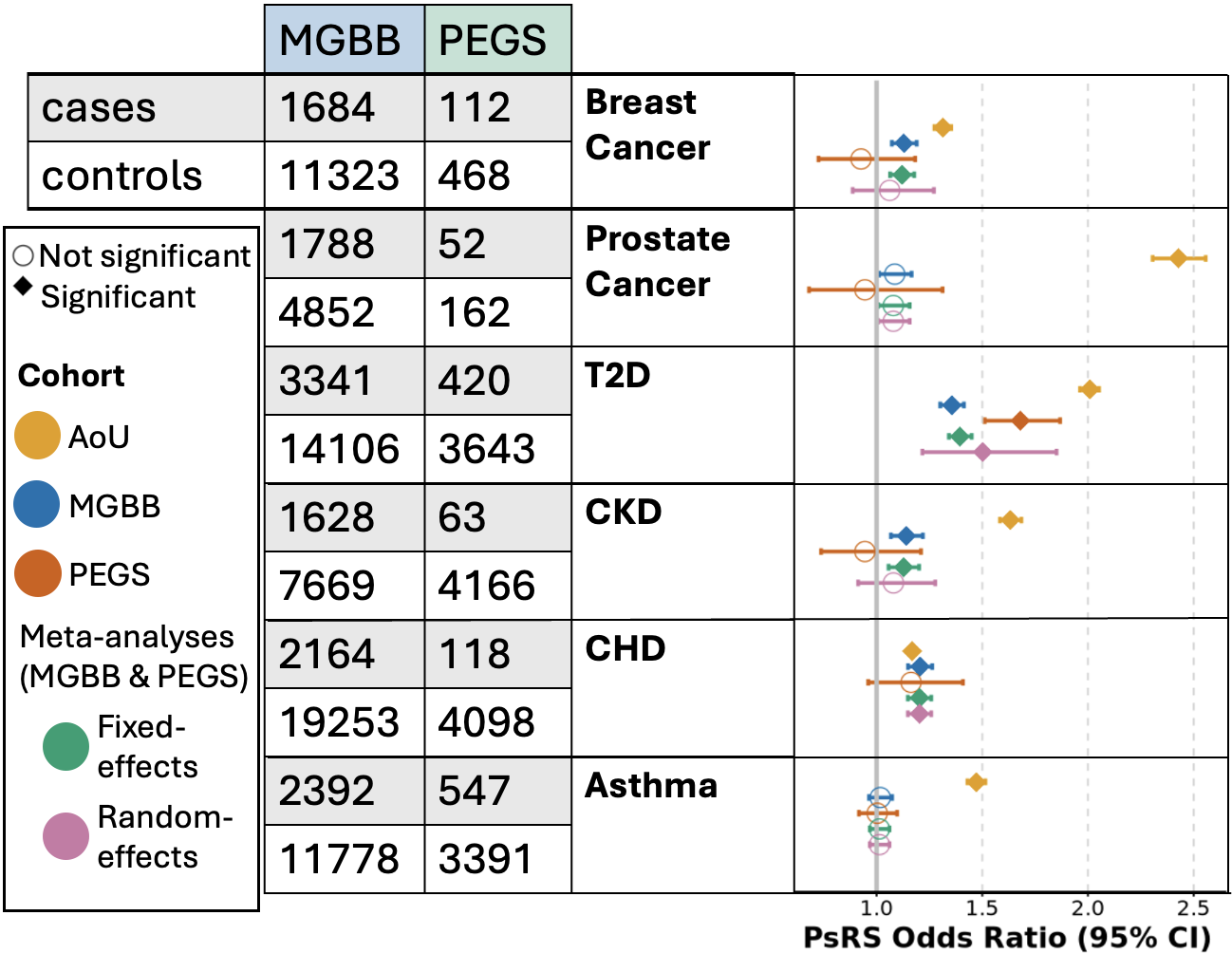

**Supplementary Figure 27:** Forest plot of PsRS OR in MGBB and PEGS, reporting both fixed and random effects meta analysis**.** PsRS OR represent the change in the odds of disease per one SD of PsRS. Numbers of cases and controls are provided. Points are color coded by cohort with fixed and random effects meta-analyses estimates in green and pink, respectively. AoU effect estimates are provided for context. Diamonds represent statistically significant effect estimates after correcting for multiple testing across 30 models (p < 1.67 × 10⁻^3^; 0.05/30).

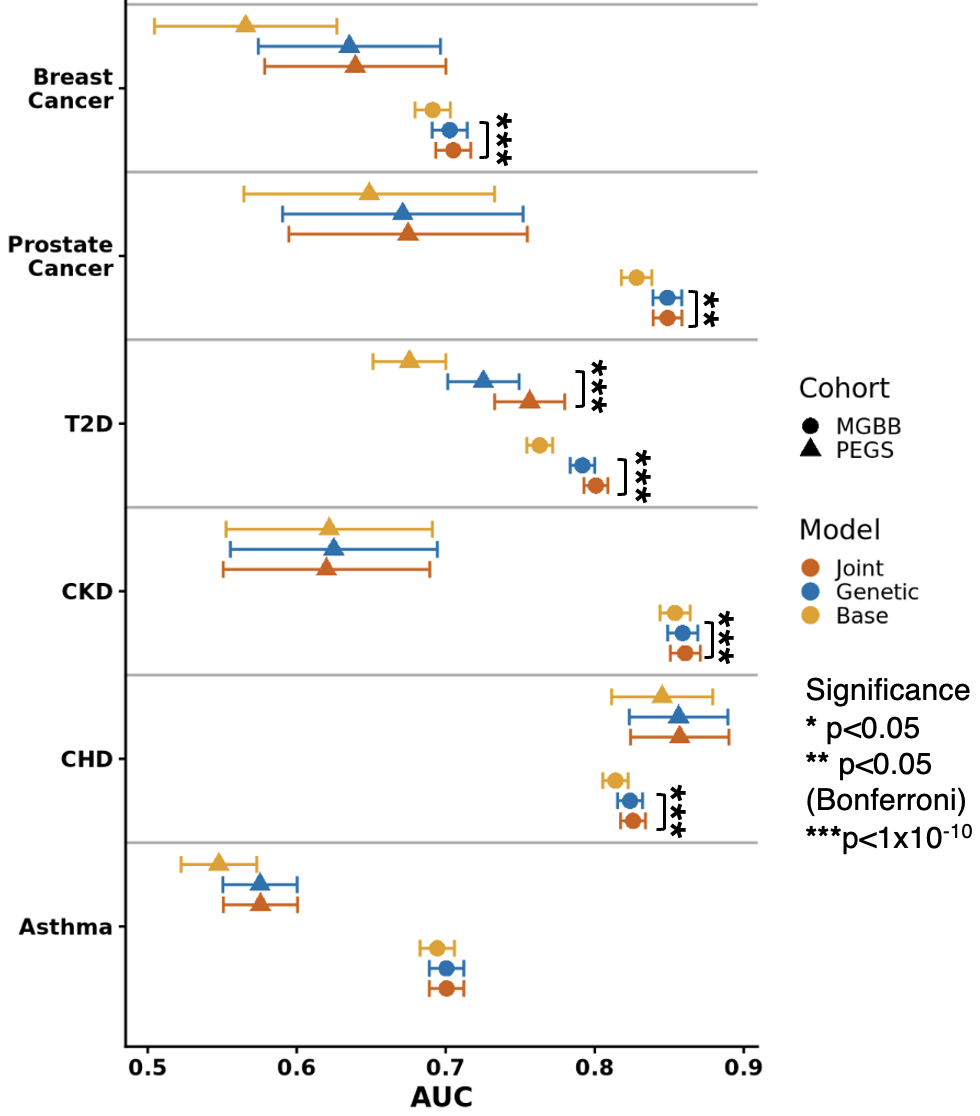

**Supplementary Figure 28:** Comparison of model discrimination (AUC) in external testing cohorts. External testing cohort is indicated by shape with MGBB as circles and PEGS as triangles. The Base model (orange) includes demographic and data availability covariates, as appropriate for each cohort. The Genetic model (blue) extends the Base model by adding PRS. The Joint model (vermillion) adds PsRS trained in AoU to the Genetic model. Significance comparisons are made between the Genetic and Joint model using DeLong’s test. One star indicates nominal significance at p<0.05, two stars Bonferroni corrected significance at p<0.05, and three stars at p<1×10⁻¹⁰.

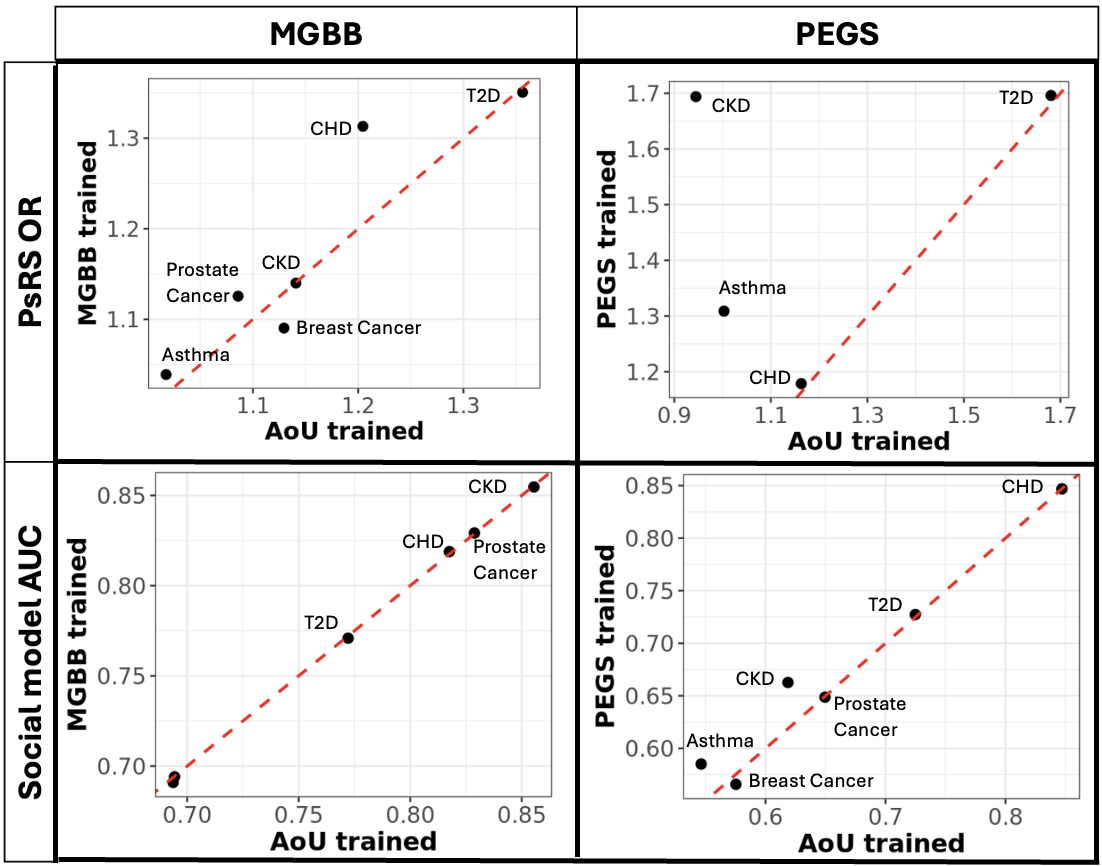

**Supplementary Figure 29:** Comparison of AoU-trained and internally trained models in MGBB and PEGS. Scatter plots compare the performance of models trained in the All of Us (AoU) cohort with models trained internally within the target cohort. The top row shows odds ratios (ORs) for poly-social risk scores (PsRS), and the bottom row shows the area under the receiver operating characteristic curve (AUC) for social models. Results for MGBB are shown in the left column, and results for PEGS are shown in the right column. The red dashed line represents the line of identity (slope = 1), where models trained in AoU and the corresponding internally trained models have identical performance. Points above the line indicate higher ORs or AUCs for the internally trained models, whereas points below the line indicate higher performance for the AoU-trained models.
